# Seasonal and Vitamin D3 mediated methylome reprogramming of peripheral immune cells and anti-viral responses

**DOI:** 10.64898/2025.12.22.25342836

**Authors:** Abhimanyu, François Lefebvre, Elisa Nemes, Nina G. Jablonski, Robert J. Wilkinson, Anna K. Coussens

## Abstract

Seasonality of inflammatory and infectious diseases is associated with several factors, including serum vitamin D fluctuations and immunological state. We longitudinally investigated peripheral blood mononuclear cell (PBMC) methylomes of healthy South African adults in summer, winter, and after high-dose winter vitamin D_3_ supplementation to determine whether vitamin D_3_ modulates seasonal epigenetic variations and whether these underpin reported seasonal transcriptional oscillations. PBMC DNA methylomes were more variable in winter compared to summer, with winter variability decreased following vitamin D_3_ supplementation. Summer and vitamin D_3_ shared differential methylation of genes involved in transcription, metabolism, and epigenome regulation. Seasonal methylome changes mapped to gene targets of viral effector proteins and pathways related to pathogenic responses, including ferroptosis. Differential methylation patterns were validated against seasonal transcriptome, accessible chromatin changes, and vitamin D receptor-independent 1,25(OH)D_3_-induced transcription in three orthogonal datasets and upon *in vitro* HIV-1 infection. These findings reveal novel epigenetic regulation of seasonal immunity and may inform new treatments for seasonal diseases.

## INTRODUCTION

Seasonality of inflammatory disease exacerbation^1,2^ and fluctuations in infectious disease presentation, including tuberculosis^3,4^ HIV-1^5^, COVID-19^6^ and influenza^7^ are well recognized. In addition to environmental factors that favor pathogen expansion and transmission, seasonal changes in disease burden are associated with oscillations in UVB atmospheric penetration. This can be due to myriads of cellular functions regulated in direct response to skin UV exposure, or due to subsequent changes in vitamin D levels (measured by serum 25-hydroxyvitamin D [25(OH)D]) induced after UVB exposure^8,9^. The frequency of circulating immune cell populations, including lymphocytes, monocytes, and neutrophils exhibit seasonal changes in immune cell transcriptomes^10^. Functionally, 5136 genes, ∼23% of the whole protein coding transcriptome of circulating immune cell populations, demonstrate seasonal fluctuations, including vitamin D receptor (*VDR*) showing increased expression in summer *vs*. winter^10^. Vitamin D and UV exposure have distinct and shared mechanisms of action on modulating immune cell functions^9^. The active metabolite of vitamin D, 1,25(OH)_2_D, activates the VDR, a transcription factor regulating >900 genes^11^. However, what governs periodicity of the seasonal transcriptome of circulating immune cells has yet to be comprehensively defined. Epigenetic mechanisms, such as chromatin accessibility, fluctuate seasonally, as observed in CD8+ T cells, regardless of age^12^. These accessibility shifts correlate with CD8+ T cell transcriptional activity^13^. In epithelial cells, *in vitro*, treatment with 1,25(OH)_2_D_3_ revealed that VDR interacts with proteins involved in DNA methylation, chromatin remodeling, and gene silencing, supporting the hypothesis that seasonal vitamin D fluctuations may drive epigenetic changes^14^.

We previously reported a prospective longitudinal study of healthy young adults in Cape Town, South Africa, where UVB radiation varies seasonally. Phlebotomy was timed to capture seasonal vitamin D_3_ peak and trough concentrations, six weeks after the summer (February-March) and winter (August-September) solstices. Participants then received six weekly oral doses of 50,000 IU vitamin D_3_ (cholecalciferol) with a third blood draw one week after the final dose (September-October). We found winter vitamin D_3_ deficiency, linked to low UVB exposure, was reversed by the regimen yielding optimal serum 25(OH)D_3_ levels post-supplementation (median >100nM)^15^. Remarkably, we identified seasonal functional differences in PBMC of these participants with vitamin D deficiency in winter, correlating with higher HIV-1 productive infection *ex-vivo*, which reversed to summer-equivalent levels following oral vitamin D_3_^15^. Given the seasonal fluctuations in PBMC transcriptome^10^ and the broad transcriptional activity of VDR^11^, we hypothesized that the seasonal and vitamin D-mediated functional differences in PBMC could be epigenetically controlled through differential DNA methylation.

Here, we report the identification of a seasonal methylome in PBMC from these individuals. Longitudinal genome-wide DNA methylation changes revealed a more variable methylome in winter compared to summer. We demonstrate that high-dose oral vitamin D_3_ partially mitigates these seasonal changes by altering methylation of genes crucial for transcription, metabolism, epigenetic regulation, and anti-viral responses, while also exerting season-independent effects. Our findings, confirmed across multiple orthogonal datasets and *in vitro* experiments, show that the seasonal immune potential of PBMC is influenced through regulation of DNA methylation, and that sustained long-term vitamin D sufficiency through supplementation may enhance anti-viral immunity in individuals at risk of winter deficiency.

## RESULTS

### Peripheral blood methylome changes between summer, winter and following oral vitamin D_3_

To investigate the hypothesis that immune cell DNA methylation patterns fluctuate by season and whether seasonal changes in serum vitamin D_3_ levels contribute to this modification, DNA was examined for 90 PBMC from 30 healthy young adults aged 19-24 years, 43.3% female. Blood was collected from each individual in Summer (S), Winter (W), and after six-weeks of Winter oral vitamin D_3_ supplementation (Winter+D_3_ [D])^15^, with timepoint mean serum 25(OH)D_3_ levels of (S) 71.3 mol/L, (W) 44.8 nmol/L and (D) 123.3 nmol/L, corresponding to sufficient, deficient and optimal levels^16^, respectively (**Fig. 1a**, **Suppl. Table S1**). As circulating immune cell frequencies can differentially vary across the year^10^, we first confirmed there were no significant difference in blood frequencies of major cell populations between timepoints for the individuals who had PBMC samples stored for methylation analysis (**Suppl. Fig. 1**).

**Figure 1.**
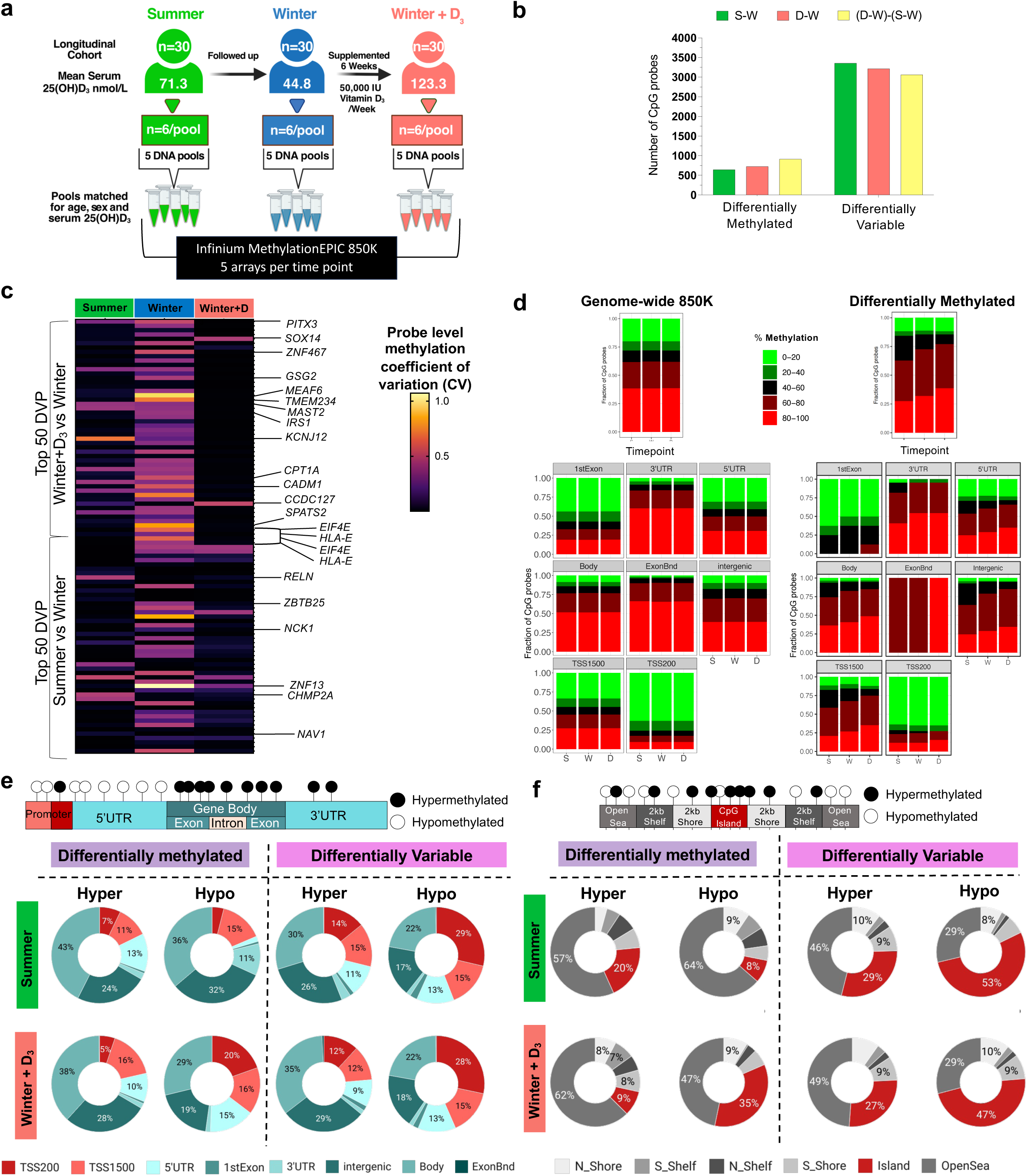
Cohort design and longitudinal methylome characteristics of peripheral blood cells. **a)** Overview of the study cohort and study design; **b)** The number of differentially methylated probes (DMP) (p<0.01) and differentially variable probes (DVP) (p<0.001) identified for each timepoint (summer (S) winter (W) and Winter+D_3_ (D)) comparison, F-test q<0.25; **c)** The coefficient of variation (CV) of top 50 most differentially variable probes (DVP) identified for Summer and for Winter+D_3_, compared to Winter (*EIF4E* and *HLA-E have DVP* in the top 50 for both comparisons, see Table S4); **d)** Methylation proportion and genomic region distribution of the 1099 DMP and all 866,659 genome-wide probes; **e)** Genomic distribution and direction of change, compared to winter, of the DMP and DVP. The donut plots are shown as a percentage of the total numbers shown in **b.** The shades of red depict the TSS, and which is the where most changes map to after the gene body**; f)** CpG loci distribution among the differentially methylated and variable regions. The Shades of red indicate the CpG islands, and a lot of differential changes localize increasingly to the CpG islands. Abbreviations: TSS, transcription start site; exonbnd, exon boundary; UTR, untranslated region.

To analyze longitudinal PBMC DNA methylation changes, the 30 DNA samples at each time point were allocated into five donor pools (n=6/pool)(**Fig 1a**), matched for age, sex, and time-point mean serum 25(OH)D_3_ levels (**Suppl. Table 1**). Each Pool was analyzed using Infinium MethylationEPIC 850K arrays to assess genome-wide DNA methylation at single CpG resolution. Following data acquisition and normalization, 790,573 CpG probes showed >0.1 standard deviation (SD) in their methylation (M)-value (logit-transformed methylation ratios) across arrays. Principal component analysis (PCA) showed clustering first based on donor DNA pool, and then timepoint (**Suppl. Fig S2a**). Normalizing for donor pool and filtering to a stricter SD >log_2_ 1.25, the resultant 60,983 probes clustered predominantly by timepoint, with Summer and Winter samples separating strongest along principal component 1 (PC1) and Winter+D_3_ samples along PC2 (**Suppl. Fig. S2b**). Comparing the β-value distributions (0-100% methylation) of all 866,659 normalized probes, Summer and Winter+D_3_ both differed significantly compared to Winter and moderately from each other (**Suppl. Table S3**).

Performing differential methylation analysis on the 790,573 probes with SD >0.1, comparing differences between the three timepoints identified 1099 top differentially methylated probes (DMP): 647 for Summer and 725 for Winter+D_3_, compared to Winter, and 914 between these two (**Fig. 1b, Suppl. Table S4**). Given the considerable impact of pool variation on PCA clustering, and the unexpectedly higher number of DMPs identified for Summer vs. Winter+D_3_, in contrast to each of their comparisons to Winter, we also performed differential variability analysis between the three timepoints to determine if sample variability was potentially impacting the number of significant DMP. This identified, compared to Winter, 3358 top differentially variable probes (DVP) for Summer, 3217 for Winter+D_3,_ and 3065 DVP between these two contrasts (**Fig. 1b, Suppl. Table S5-7**). Indeed, analysis of the coefficient of variation (CV) of methylation β-values for the top 50 DVPs for both the S-W and D-W contrasts showed most probes were hypervariable in Winter with 54% of the top 100 DVPs showing a CV >30% in winter but only 14% and 7% in Summer and Winter+D_3_, respectively (**Fig. 1c, Suppl. Table S8**). Increased variability in DNA methylation in Winter samples, thus explained the lower number of top significant DMPs identified when Summer and Winter+D_3_ samples were compared to Winter.

### Methylation changes associated with Summer and Vitamin D_3_ supplementation are enriched in gene regulatory regions and CpG Islands

Assessing percentage methylation (β-value) distributions of the top 1099 DMP in comparison to the 866,659 genome-wide probes (**Fig. 1d**), showed DMP were least abundant in probes with 0-40% methylation and most frequently intermediately methylated (40-80%). The lower proportion of 0-20% methylated probes was most notable within promoter regions (200bp of the transcription start site [TSS200]), in comparison to a larger proportion in the 1st Exon. Overall, Winter+D_3_ showed a greater proportion of 60-100% methylated probes across most genomic locations, except the 1st exon.

Examining the potential gene-regulatory effects of Summer and Winter+D_3_ DMP and DVP, compared to Winter, we stratified these differences by direction of methylation change and genomic location (**Fig. 1e**, **Supp**l. **Fig. S2c** and **Supp**l. **Table S9**). Hyper– and hypomethylated DVP for both Summer and Winter+D_3_ were significantly overrepresented in the promoter regions (TSS200). They also shared overrepresentation of hypomethylated DVP in the TSS1500, 5’UTR and 1st Exon and underrepresentation in the 3’UTR, intergenic, gene body, and exon boundaries; a methylation pattern suggesting associated DNA would be more accessible for transcription factor binding. The only consistent opposing pattern was hypomethylated DMP in the TSS200, being underrepresented in Summer but overrepresented in Winter+D_3_, with hypermethylated DMP in TSS200 also significantly underrepresented in Winter+D_3_. When comparing the location of CpGs based on island, shore, shelf or open sea (**Fig. 1f** and **Suppl. Table S9**) DMP in CpG islands were the most significantly enriched for Summer and Winter+D_3_ (under and over represented, respectively) and both showed overrepresentation of hypermethylated DVP in CpG islands; further supporting methylation differences associated with Summer and Winter+D_3_ are in functional regions of the genome,.

### Summer and Winter Vitamin D_3_ supplementation induced shared and unique differential DNA methylation in PBMC

Of the 647 top Summer DMP 506 were hypomethylated and 141 hypermethylated, compared to Winter, whereas for the 725 top Winter+D_3_ DMP, 184 were hypomethylated and 541 hypermethylated (**Fig. 2a-b**). Consistent with Winter contributing the great variability between samples, the number and direction of DVP, compared to Winter, were extremely similar for Summer (2568 Hyper and 790 Hypo) and Winter+D_3_ (2547 Hypo and 669 Hyper) (**Suppl. Fig. S3a-b**). 297 DMP were shared between Summer and Winter+D_3_ (46% and 41% of each, respectively) and 825 top DVP were shared (27% and 28%, respectively) (**Suppl. Fig. 3c)**. Analyzing DMP based on those annotated to genes or intergenic locations, summer had 442 differentially methylated genes and 196 intergenic DMP whilst Winter+D_3_ had 519 differentially methylated genes and 188 intergenic DMP, 307 in total shared (48% and 43%, respectively) (**Fig. 2c).** Among gene-annotated DMP, 84 were commonly hypomethylated and 50 commonly hypermethylated in Summer and Winter+D_3_, and for DVPs the numbers were 559 and 117, respectively (**Suppl. Fig. S3d** and **Suppl. Table S10**). Thus, overall, Summer and Winter+D_3_ shared more commonly hypomethylation, compared to Winter.

**Figure 2.**
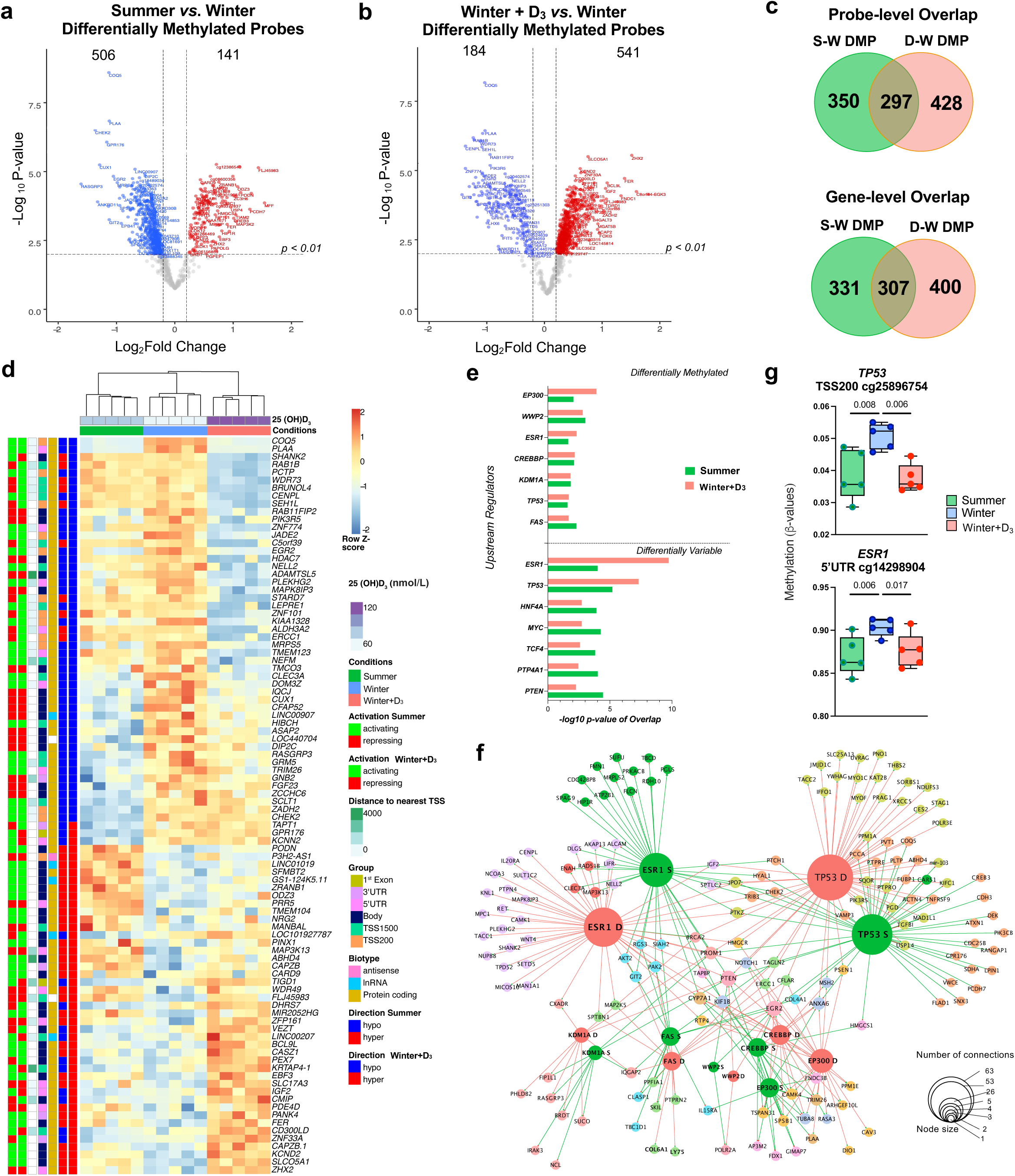
Season and oral Vitamin D_3_ induce widespread shared and unique differential methylation in PBMCs. Volcano plots showing hyper-(red dots) and hypo-(blue dots) differentially methylated probes (DMP) compared to winter, in **(a)** Summer and in **(b)** Winter+D_3_ supplementation; **c)** Venn showing overlap between Summer compared to Winter (S-W) and Winter+D_3_ compared to Winter (D-W) DMP based on probe ID (probe-level) or gene/intergenic annotation (gene-level); **d)** Heatmap of top 50 DMP each for S-W and D-W comparisons. Methylation beta-values have been plotted as a normalized z-score with red showing increased methylation and blue lower methylation. Red and blue blocks per row on the left show the hyper– and hypo-methylation status as compared to winter. Predicted activation depending on the genomic location is shown in red (repressed) and green (activated). Gene biotype and the location is depicted under the label “group”; **e)** Summary of predicted top regulators as per Ingenuity pathway analysis (see Suppl. Fig. S3e-g); **f)** Network depiction of results for (**e**), with each gene with DMP connected to its predicted upstream regulator through a line the color of which is defined by whether the gene connected to a summer (green) or Winter+D_3_ (red) DMP. The size of the node is proportional to the number of connections it makes with the proteins in the network. The smallest node size means one connection and the ‘hub’ genes are regulators such as *TP53*, *ESR1*,*CREBBP*,*EP300* etc. The legend is proportional to the node size shows the number of connections; **g**) *TP53* and *ESR1* show highest methylation in winter samples. Each dot corresponds to one DNA pool, line median, p-values from DMP F-test.

Among the DMP, the top two with greatest significance for both Summer and Winter+D_3_ were in genes *COQ5* and *PLAA,* both hypomethylated, compared to Winter (**Fig. 2a, b, d**). The shared and unique nature of the top 50 DMP for Summer and Winter+D_3_ is seen in the gene-level methylation heatmap (**Fig. 2d**), where their generally common direction of gene regulation, either activation or repression, based on the probe methylation status and genomic location (as defined by Paul et al.^17^) is demonstrated.

Next, predicted upstream regulators were analyzed in both sets of DMP and DVP using Ingenuity Pathway Analysis (IPA), identifying a large overlap in the top regulators of genes within the Summer and Winter+D_3_ methylome (**Fig. 2e-g**, **Suppl. Fig. S3e-g**). Network-based analysis and representation indicated that *TP53* and *ESR1* were the top shared common gene regulators, showing most network connections with both the Summer and Winter+D_3_ DMP and DVP genes, as indicated by the size of their network nodes (**Fig. 2f)**. Other important and shared regulator genes included *EP300*, *WWP2*, *CREBBP*, *KDM1A* and *FAS* (**Fig. 2e-f**), many of these top regulators controlling epigenome changes^18,19^, including DNA methylation. The top two predicted regulators *TP53* and *ESR1* were both found to be hypomethylated in Summer and in Winter+D_3_ compared to Winter hypermethylation (**Fig. 2g**).

### Metabolic and Epigenetic pathway genes are predominately hypomethylated in Summer and by Vitamin D_3_

To gain greater insights into the potential functional impact of differential gene methylation, we carried out multiple pathway analyses on DMP and DVP. Gene ontology (GO) enrichment analysis identified several processes that were perturbed with change in methylation between seasons and Winter+D_3_ (**Fig. 3a and Suppl. Table S11**). To summarize the enriched GO output for interpretability, we employed *Enrichment map*^20^ in Cytoscape^21^ (**Suppl. Fig. S4)**. Enrichment map connects GO processes with shared genes and determines the most accurate enrichment annotations. A total of 49 enriched GO processes encompassing DMP and DVP were identified, which were inferred into ten broad categories (**Suppl. Table S11)**. Of the enriched processes, 6% (3/49) were shared by DMP and DVP in Summer (S-W), and 20% (10/49) for Winter+D_3_ (D-W) (**Fig. 3a**). The two largest GO inferred pathway categories were metabolic processes and epigenetic regulators, followed by transcription/translation, immune function, transmembrane transport, cell death, phospholipid homeostasis, aging, and response to pathogen. Cell death pathways included apoptosis, UV protection and response to vitamin D. Among the commonly enriched metabolic processes between Summer and Winter+D_3_ were insulin receptor signaling, glutamate cysteine activity and fatty acid metabolism, with shared epigenetic processes including condensed chromosome kinetochore that had the most significant enrichment of all DMP enriched terms (**Fig. 3a and Suppl. Table S11).**

**Figure 3.**
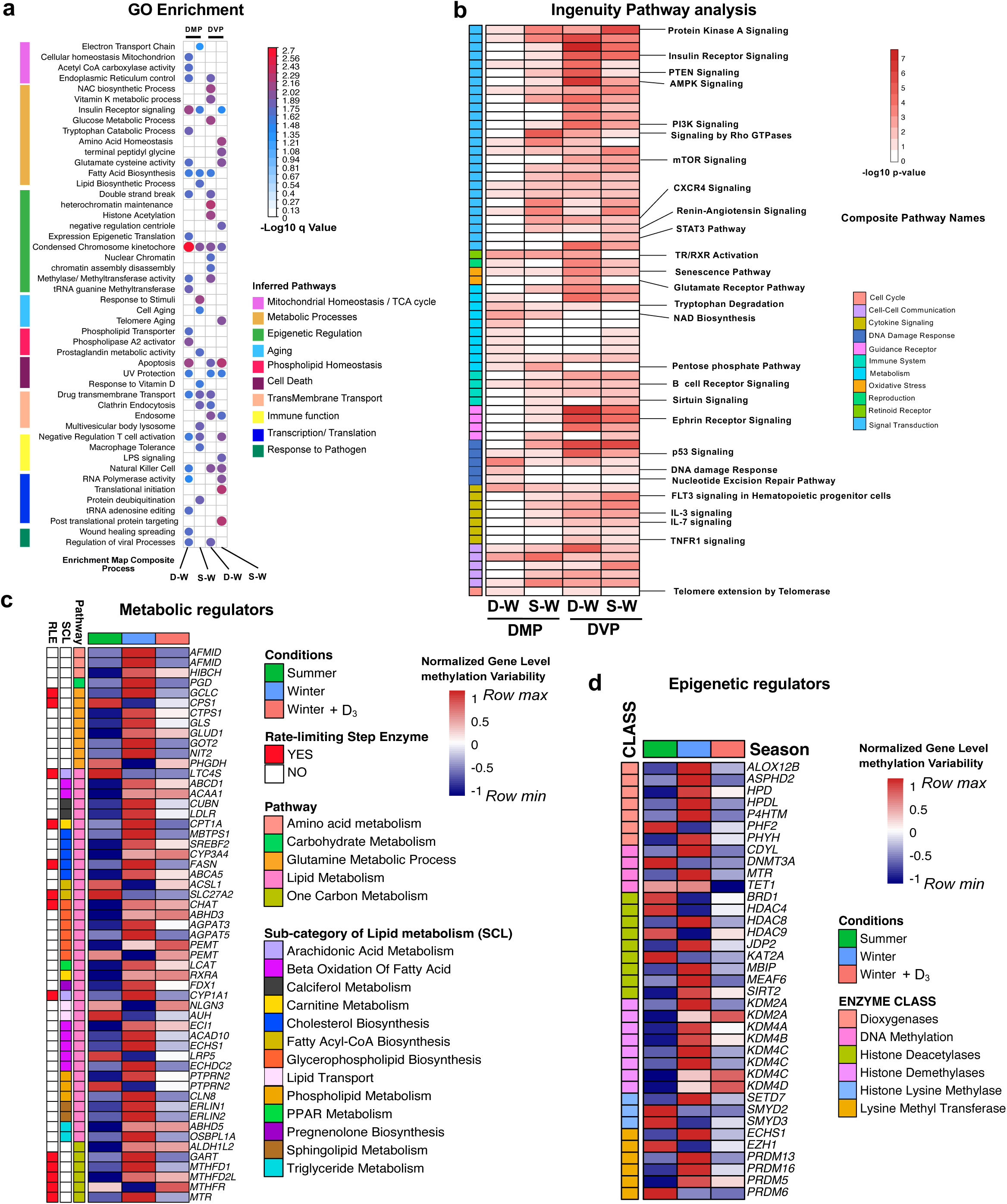
Shared and unique functional pathway enrichment in Summer– and Vitamin D-related methylome changes. **(a)** Composite plot derived from Gene Ontology (GO) and Enrichment map analysis (in Suppl. Fig. S3) of differentially methylated probes (DMP) and differentially variable probes (DVP) for Summer vs Winter (S-W) and Winter+D_3_ vs Winter (D-W), showing metabolic process enrichment, the colors represent the –log10 q-values; Similar enrichment mapped using Ingenuity Pathway analysis **(b)**, the colors represent –log10 p-values. Empty squares or white color shows that the corresponding comparison was not enriched for that process; DMPs and DVPs (p < 0.01 0.001, respectively) were used for the enrichment. Common themes were identified using ‘shared’ Reactome terms. **c)** Metabolic regulators as identified by pathway analyses presented in (a). Normalized gene level methylation variability (NGV) is plotted. High NGV represents higher levels of methylation variability. Three levels of metabolic annotations are plotted. The overall metabolic class, whether the regulator is a rate-limiting step (RLE) enzyme as defined by Zhao et al.^23^. and the metabolic sub class for Lipid metabolism genes, derived from Reactome and confirmed with other databases when necessary; **d)** Epigenetic enzyme as identified by the pathway analysis in (a). The enzyme class derived from Reactome is presented as row annotations and NGV is plotted.

IPA enrichment generated 66 pathways that were significant for at least one differential comparison. Interestingly, both the DMP and DVP were enriched in remarkably similar IPA processes up to 90% (60/66) for Summer and 75% (50/66) for Winter+ D_3_ (**Fig. 3b and Suppl. Table S12-13)**. Deriving the IPA pathways composite terms from Reactome, Signal transduction (including PI3K, mTOR, PTEN, AMPK, CXCR4 and IGF-1 signaling), Metabolism (including NAD biosynthesis, Pentose Phosphate Pathway [Oxidative Branch] and Pyridoxal 5’-phosphate Salvage Pathway), DNA damage response (including p53 signaling), cytokine signaling (including FLT3, IL-3, IL-7, and TNFR1 signaling) and immune signaling (including Role of NFAT, B Cell Receptor Signaling and Fcγ Receptor-mediated phagocytosis in macrophages and monocytes) were the top five largest composite enriched terms shared by Summer and Winter+ D_3_ (**Fig. 3b and Suppl. Table S12**). Interestingly, metabolic pathways NAD biosynthesis, and Salvage Pathway are responsible for epigenetic changes and involved in DNA methylation^22^. Vitamin D signaling is known to interact with retinoid X receptor (RXR) signaling^14,22^ and notably TX/RXR activation was shared in enriched DMP in Summer and Winter+D_3_ (**Fig. 3b**). Interestingly, glutamate pathway enrichment was common to both GO and IPA, indicating importance of this pathway in the differential methylation changes and *GOT2,* a key regulator of glutamine and tryptophan metabolism was hypervariable in winter (**Fig. 3c**).

Next, we investigated the DNA methylation variability among genes of the top two GO enriched pathways, Metabolic (**Fig. 3c**) and Epigenetic (**Fig. 3d**) regulators. This identified that the important regulators of metabolic pathways were hypervariable, with increased gene level methylation variability in Winter samples as compared to Summer or Winter+ D_3_ (**Fig. 3c**). Rate limiting step enzymes (RLE), enzymes that guide metabolic processes^23^, such as Glutamine metabolism (*GCLC*, *CPS1*), Lipid Metabolism (*LTC4S*, *CPT1A*, *FASN*, *SLC27A2*, *CHAT*, *CYP1A1*) and one-carbon metabolism (*GART*, *MTHFD1*, *MTHFD2L*, *MTHFR*, *MTR*) were hypervariable in Winter and commonly hypovariable in Summer and Winter+D_3_ (**Suppl. Table S13**). Since lipid metabolism was the most represented enriched metabolic process, a sub-classification using Reactome database of lipid metabolism pathways highlighted the importance of lipid metabolic processes that regulate the inflammatory response that included arachidonic metabolism, glycerophospholipid metabolism, and phospholipid metabolism (**Fig. 3b**). For epigenetic regulator, we also found a similar hypervariable DNA methylation in Winter including for key enzymes classes, dioxygenases such as *ALOX12B*, *HPD* and critical DNA methylating enzymes *DNMT3A* and *TET1* (**Fig. 3d**). Other epigenetic processes included histone modifiers such as histone deacetylases (including *HDAC4*, *HDAC8*, *HDAC9, KAT2A*, *SIRT2*), demethylases (*KDM4A, KDM4B, KDM4C, KDM4D*), lysine methylases (*SETD7*, *SMYD2*, *SMYD3)* and lysine methyl transferases (including *PRDM5, PRDM13, PRDM16*). Together, these data demonstrate a widespread dysregulation of DNA methylation in key metabolic and epigenetic regulators in winter, with variability between individuals generally reduced in summer and after vitamin D_3_ supplementation in winter.

### Genes in key pathways involved in HIV-1 and SARS-CoV-2 regulation are hypermethylated in winter and show direct interaction with viral proteins

To reduce pathway results dominated by genes with multiple CpGs, we next explored only the DMP for further pathway analysis using the Human Molecular Signatures Database MsigDB-C2^24^ and KEEG^25^ with an over-representation test where the length of the gene was considered. Only DMP were investigated since DMP and DVP showed shared pathway level similarity and winter hypermethylation in previous analyses. Of the resultant pathways, similar pathways were merged into common composite terms based on if they shared the same Reactome “has” term and based on published literature (**Suppl. Table S14**). This revealed common pathway methylome themes, with the largest gene enrichment in Winter+D_3_ methylome also over-represented in the Summer methylome, leading with genes involved in PI3K-AKT signaling, gene transcription, DNA replication and cell cycle regulation (**Fig. 4a-b**). To understand the direction of changes at a pathway level, we derived a summed Z-Score of methylation β-values, which incorporates the methylation status of each gene in the pathway. Pathway-level methylation score showed a similar pattern across pathways with the highest methylation in Winter, that was reduced in Winter+D_3_ and in Summer (**Fig. 4c**), consistent with observations for GO and IPA analyses.

**Figure 4.**
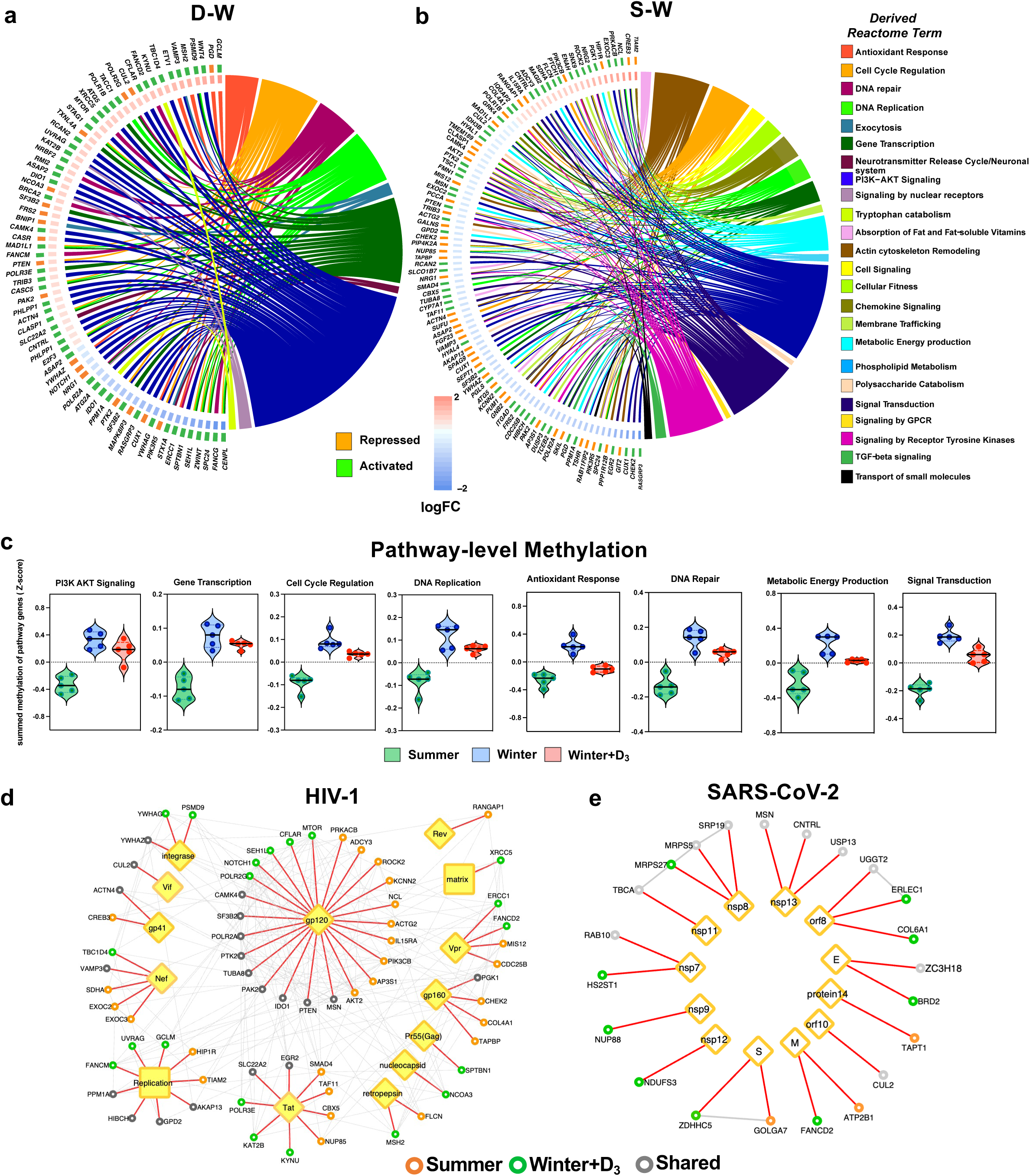
Common and unique pathway level changes in summer and vitamin D methylome dominated by PI3K signaling, DNA replication/repair and antiviral responses. Chord plots showing shared genes among multiple pathways which are commonly over-represented with differentially methylated probes (DMP) considering gene length, in Summer **(a)** and Winter+D_3_ **(b)**. DMPs p < 0.01 were used for enrichment analysis from MsigDB-C2 and KEEG. Common themes were identified using shared Reactome terms. Each gene has it’s log_2_ fold change (FC) as compared to Winter shown and each gene also has its predicted activation shown, Orange (Repressed) and green (activated); **c)** Summed Z-score of the four common pathways between Summer and Winter+D_3_ and two top pathways for Summer and for Winter+D_3_ enrichment; **d**-**e)** Protein-protein Interaction (PPI) network for genes containing DMPs with known viral proteins interactions for (**d**) HIV-1, derived from the HIV-1-human interaction database^28^ and (**e**) SARS-CoV-2 from Gordon et al.^32^. Direct enriched interaction is shown with red connected lines, grey lines show PPI between the human proteins. Gene circles indicate if the gene contains DMP identified in Summer (orange circle), Winter+D_3_ (green circle), or shared by both (grey circle), compared to winter.

Since PI3K-AKT signaling was the top shared pathway for Summer and Winter+D_3_, demonstrating relative winter hypermethylation and PI3K-AKT signaling is known to be particularly important for HIV-1 control^26,27^. Given our previous work demonstrating a winter associated increase in *ex vivo* HIV-1 replication in PBMC from these same individuals, we next investigated whether genes with DMP were directly involved in HIV-1 control. Mapping all the genes with top DMP from either the Summer or Winter+D_3_ comparison (**Fig. 2a-b, Suppl. Table S4)** to the NIH Human and HIV-1 interactome database^28^, we identified 65 differentially methylated genes that have direct interaction with a total of 14 HIV-1 proteins, the largest interacting nodes being for HIV-1 gp120 and Tat, interacting with 24 and 9 genes, respectively (**Fig. 4d**). To determine if these methylome changes could affect immune responses to other seasonal viruses, given the proposed role of vitamin D deficiency on SARS-CoV-2 severity^29–31^, we performed a similar analysis for SARS-CoV-2 protein interactions using the SARS-CoV-2-human interactome derived from Gordon et al.^32^ (**Fig. 4e**). This identified 22 genes that have direct interaction with a total of 12 SARS-CoV-2 proteins. Comparing common gene interactions between the two viruses identified three-anti-viral immunity genes *CUL2*, *MSN*, and *FANCD2* common to both HIV-1 and SARS-CoV-2 human protein interactions. Together, these analyses indicate that methylation changes that occur in summer, and those induced by vitamin D_3_ are enriched in functional pathways important for anti-viral responses.

### Seasonal epigenome changes concord with transcriptional and chromatin changes across diverse cohorts

To validate that observed methylation changes translate to seasonal transcriptional oscillation, we next investigated the seasonal transcriptional profiles of genes with Summer and Winter+D_3_ DMP within the publicly available seasonal transcriptome dataset by Dopico et al.^10^. Using weighted gene correlation network analysis (WGCNA) they defined modules of co-regulated genes with season-specific oscillations across globally distributed populations, including VDR and ESR summer increased expression. Seven seasonal modules were defined as winter high expression and three as summer high expression (**Suppl. Table S15**)^10^. Gene-set enrichment analysis (GSEA)^33^ was used to compare the enrichment of genes with DMP in Summer and Winter+D_3_ to genes within the 10 seasonal transcriptome modules, with the resultant normalized enrichment score (NES) indicating the directionality of methylation enrichment. Genes in the seven winter high modules were positively enriched (ie. hypermethylated) in summer DMP, whilst genes in the three summer high modules were negatively enriched (i.e. hypomethylated) in summer DMP (**Fig. 5a**); consistent with a decrease in DNA methylation facilitating an increase in expression, and vice-versa^34^. The Winter+D_3_ DMP also showed concordant directional enrichment for 3 winter and 1 summer high module genes, but also showed inverse enrichment or 3 winter and 2 simmer high modules (**Fig. 5a**), reflecting both seasonal-associated and –independent methylation by vitamin D_3_. Plotting the methylation β-values of the top 100 enriched seasonal module genes, further demonstrates Summer and Winter+D3 share a consistent effect on seasonal-associated DMP (**Fig. 5b**). Many of these genes are related to immune exhaustion and other critical immune processes, including the top two DMP for both comparisons, *PLAA* and *COQ5*.

**Figure 5.**
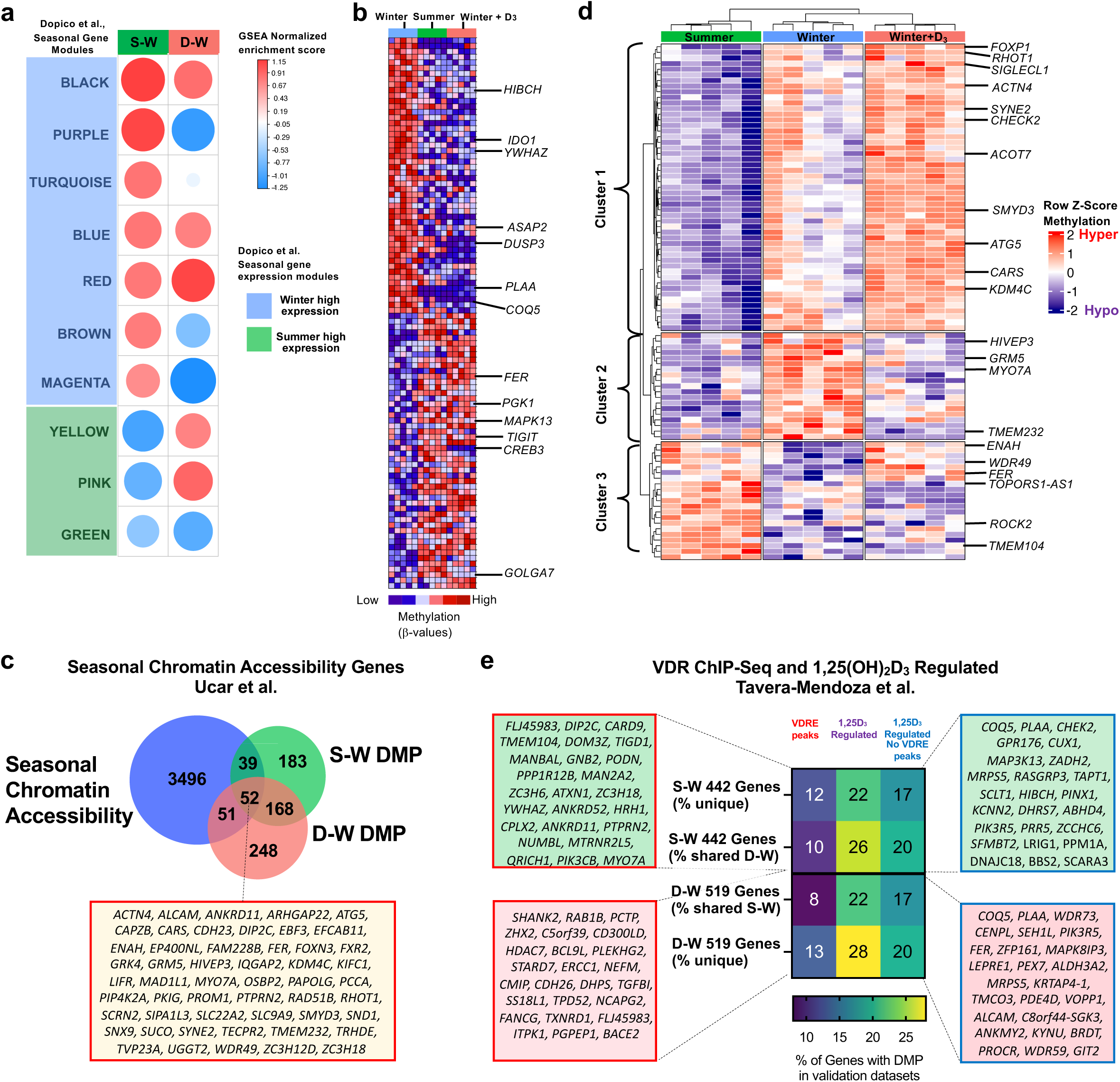
Identified seasonal and vitamin D_3_ associated methylation changes correspond with transcriptional changes in diverse publicly available datasets. **a**) Enrichment of genes with differentially methylated probes (DMP) in genes within the summer and winter high gene expression modules identified by Dopico et al.^10^ by weighted gene correlation network analysis (WGCNA), the colors represent the directionality hyper (red) and hypo (blue) of the normalized enrichment score (NES), and the size of the dot corresponds to the NES, denoting effect size, obtained from a GSEA analysis; **b**) Heatmap of the seasonal methylation differences (β-value) for DMP in the top 100 genes identified in Dopico et al. to be enriched in Summer vs Winter; **c**) Overlap of genes with seasonal chromatin accessibility identified by Ucar et al.^12^ and genes identified with DMP in summer (S-W) and after winter vitamin D supplementation (Winter+D_3_, D-W), compared to winter, with 52 shared genes between all three listed; **d**) Heatmap of the 91 shared genes from (**c**) with seasonal chromatin accessibility and Summer DMP separated into three clusters based on seasonal differences in methylation Z-score; **e**) Percent of genes with Summer (S-W) and Winter+D_3_ (D-W) DMP that were also identified by Tavera-Mendoza et al.^14^ to have vitamin D receptor (VDR) binding to promoter vitamin D response elements (VDRE) by ChIP-seq (VDRE peaks) in MCF-7 fibroblast cells, genes expressed in MCF-7 cells following 1,25(OH)_2_D_3_ treatment (1,25D_3_ regulated) and genes that were 1,25D_3_ regulated but did not have a VDRE peak. Percentages calculated for each DMP list for genes with DMP shared by both S-W and W-D and when not shared indicated as unique. Boxes list top 25 genes with DMP that overlap with genes with VDRE peaks (red border) and genes 1,25D_3_ regulated but did not have a VDRE peaks (blue border).

Further validating the identified seasonal methylome we next compared genes with DMP to a second publicly available dataset of 3638 genes defined in PBMC to have seasonal changes in chromatin accessibility from Ucar et.al^12^. Summer DMP overlapped with 91 genes and Winter+D_3_ DMP with 103 genes, with 52 in common (**Fig. 5c and Suppl. Table S16**). The 91 seasonal chromatin accessibility genes formed three clusters based on Summer DMP methylation values (**Fig. 5d**). Cluster 1 was hypomethylated in Summer with variable hypermethylation in Winter and consistent hypermethylation in Winter+D_3_. Genes included *FOXP1, SIGLECL1, CHECK2, ACOT7, SMYD3, ATG5, CARS,* and *KDM4C*, performing processes such as autophagy, ferroptosis and epigenetic regulation. Cluster 2 was hypomethylated in Summer and Winter+D_3_ and Winter hypermethylated, included genes such as *HIVEP3*, *GRM5*, *MYO7A* and *TMEM232*. Cluster 3 represented Summer hypermethylated genes that were relatively hypomethylated in Winter and either hyper or hypomethylated in Winter+D_3_, including *FER*, *ROCK2*, *TMEM104*, and *TOPORS1-AS1* (**Fig. 5d**).

The final validation was performed against a public dataset from a study of MCF-7 epithelial cells treated with the active vitamin D metabolite 1,25(OH)_2_D_3_ that identified 1,25(OH)_2_D_3_ regulated genes and those that contain vitamin D receptor (VDR) binding vitamin D response elements (VDRE) in their promoters by VDR Chromatin Immunoprecipitation (ChIP)-seq^14^. They reported 6036 genes with VDR-binding VDRE and 9210 genes differentially expressed after 1,25(OH)_2_D_3_ treatment. Summer and Winter+D_3_ DMP genes had 22% and 21% overlap with the VDRE containing genes, respectively, and 48% and 50% overlap with 1,25(OH)_2_D_3_ regulated genes (**Fig. 5e**). Investigating specifically the genes regulated by 1,25(OH)_2_D_3_ that lacked a VDRE for direct transcriptional activation by 1,25(OH)_2_D_3_ bound VDR could potentially indicate those epigenetically regulated. We found 37% of Winter+D_3_ and Summer DMP in these genes, with 20% representing shared genes and 17% unique to both. The top 25 DMP for both sharing *COQ5*, *PLAA*, *MRPS5* and *PIK3R5* (**Fig. 5e**). Taken together, these comparisons against three external datasets provide orthogonal validation to our discovery and show that the differentially methylated genes we identified associate with the seasonal transcriptome, seasonal chromatin accessibility, and 1,25(OH)_2_D_3_ mediated transcriptional changes dependent and independent of VDRE.

### Differential methylation of ferroptosis-associated genes translates to differential gene expression during *in vitro* HIV-1 infection

Finally, to validate one of the functional pathways perturbed by the combined effects of DMP and DVP we performed g:Profier^35^ enrichment of KEEG and WikiPathways. This identified ferroptosis, an iron-dependent cell death pathway involving ferritinophagy and lipid peroxidation, and many of the constituent pathways that contribute to this necrotic inflammatory mode of programmed cell death, including glutathione and cysteine metabolism (e.g. *GPX4, GCLC, GCLM, CBS*), eicosanoid synthesis and arachidonic acid metabolism (e.g. ALOXs), and NRF2 signaling (*NFE2L2*) (**Fig. 6a-b**) were enriched in our DMP GO and IPA analyses (**Fig. 3a-c)**. The top 2 DMP identified for both Summer and Winter+D_3_ comparisons, *COQ5* and *PLAA*, are both involved in ferroptosis. *COQ5* encodes Coenzyme Q5, a methyltransferase that catalyzes the penultimate step of COQ10 production, an important inhibitor of ferroptosis^36^. Similarly, *PLAA* encodes phospholipase A2 activating protein, an important enzyme for membrane lipid homeostasis, depletion of which sensitizes cells to ferroptosis^37,38^. Additionally, TP53 and ESR1, the identified top regulators of DMRs and DVP (**Fig. 2e-g**), are also critical regulators of ferroptosis, TP53 through its regulation of multiple involved metabolic pathways and ESR1 through its regulation of the cystine transporter SLC7A11^39,40^. Of the 31 genes identified to be directly involved in ferroptosis, 6 were differentially methylated and 25 variably methylated, the majority being hypomethylated in Summer and Winter+D_3_ (58% and 64%, respectively, relative to Winter) resulting in predicted activation based on genomic positions (**Fig. 6c**, green boxes).

**Figure 6.**
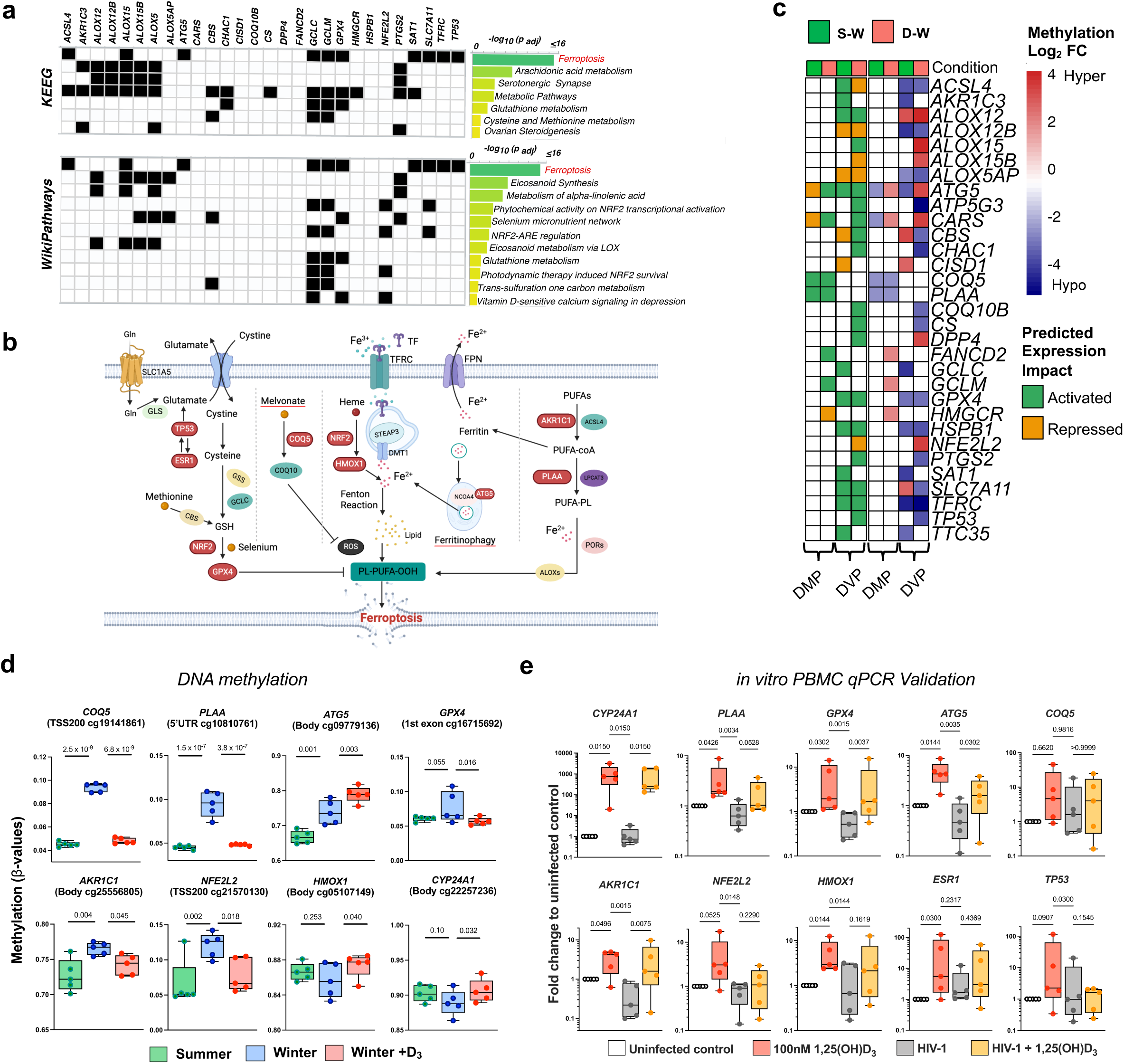
Ferroptosis related genes are differentially methylated with change of season and vitamin D_3_ supplementation attenuating HIV-1 mediated downregulation of ferroptosis regulators. **a)** g:Profiler identifies ferroptosis as one of the major perturbed pathways. All probes with differential methylation F-Test p<0.05 were used in the analysis. KEEG and WikiPathways annotations are shown. Black = present, White = absent;); **b**) Schematic of ferroptosis signaling pathways, with genes identified to have DMP and/or DVP in red bubbles, made in Biorender. **c**) Heatmap of the ferroptosis genes enriched from KEEG in the various differential lists. On the left are annotations for predicted gene expression activation (green) or repression (gold), and on right, log_2_ fold change (FC) of each DMP and DVP comparing summer to winter (S-W) or following 6-weeks winter vitamin D_3_ supplementation to winter (D-W). **d)** Methylation level (β-value) of the most significant probe (pairwise p-values from the differential methylation F-test) for seven genes involved in ferroptosis signaling (see b) and *CYP24A1* at each time point. Each dot corresponds to one DNA pool. *COQ5*, *PLAA*, *ATG5*, were top DMP; *HMOX1*, *GPX4*, *NFE2L2*, *AKR1C1*, CYP24A1 q<0.51. **e)** Relative expression of ferroptosis related genes and *CYP24A1* (positive control, transcriptionally induced by 1,25(OH)_2_D_3_) in PBMC from healthy donors (n=5) 9 days post *ex vivo* HIV-1 infection, cultured with or without 100nM 1,25(OH)_2_D_3,_ compared to uninfected PBMC. Quantitative realtime RT-PCR (pPCR) was calculated using 2^-ddCT, with each gene normalized to housekeeping gene *RPL13A.* Indicated q-value from Freidman test with Benjamini, Krieger and Yekutieli False discovery rate.

Given the role of ferroptosis in HIV-1-mediated immune cell death^41^ and our prior finding infecting PBMC from these same participants withHIV-1 that winter oral vitamin D_3_ reduced the winter-associated increase in HIV-1 replication *ex vivo*^15^, we next tested whether *in vitro* 1,25(OH)_2_D_3_ treatment of PBMC results in transcriptional modulation of selected ferroptosis-related genes and whether such 1,25(OH)_2_D_3_ modulation can also occur during HIV-1 infection. PBMC isolated from five new healthy donors in winter were infected with HIV-1 and then cultured for 9 days in the absence or presence of 100nM 1,25(OH)_2_D_3_, with gene expression at day 9 compared to uninfected untreated controls. We measured transcriptional changes in *PLAA*, *COQ5*, *TP53*, *ESR1* and additional selected key regulators within the ferroptosis pathway including those on DVP or DMP lists: *GPX4* (ferroptosis inhibitor), *NFEFL2* (encoding NRF2), *ATG5* (ferritinophagy regulator)*, AKR1C1* (negative regulator of ferroptosis induced by NRF2^42^) and HMOX1 (heme oxygenase). Plotting the β-value of the probe with the most significant pairwise comparison from the differential methylation analysis for each gene showed Winter vitamin D_3_ generally having an activating effect, compared to Winter (noting not all were top DMP) (**Fig. 2g and 6d**). Additionally, *CYP24A1* was measured as a positive 1,25(OH)_2_D_3_ transcriptional regulation control, given it is highly induced by 1,25(OH)_2_D_3_*-*mediated VDRE activation^14^. Consistent with this, 1,25(OH)_2_D_3_ significantly induced *CYP24A1* expression >100-fold in HIV-1 infected and uninfected PBMC. Following a similar pattern of induction but with a median increase <10-fold 1,25(OH)_2_D_3_ increased expression of *PLAA*, *GPX4*, *ATG5*, *AKR1C1, ESR1*, *HMOX1* and *NFE2L2* in uninfected PBMC. During HIV-1 infection 1,25(OH)_2_D_3_ also increased expression of *PLAA*, *GPX4*, *ATG5*, and *AKR1C1*, but the effect on *ESR1*, *HMOX1* and *NFE2L2* was perturbed (**Fig. 6e**). *TP53* and *COQ5* showed similar but non-significant trends with greater variability in donor responses. Together, this data confirms that genes with identified seasonal and vitamin D_3_ modulated DNA methylation changes within the ferroptosis pathway are transcriptionally modified in PBMC following 1,25(OH)_2_D_3_ treatment.

## DISCUSSION

We investigated seasonal methylome-wide changes in PBMC from 30 healthy young adults exposed to seasonal UV variation and winter vitamin D_3_ supplementation. We identified that the winter methylome undergoes genome-wide changes and exists in a relatively hypermethylated and hypervariable state, compared to summer. We further demonstrated that winter vitamin D_3_ supplementation induced both similar and distinct methylome changes compared to summer. Using complementary and iterative bioinformatic and experimental approaches, we observed multiple levels of regulation based on the differential methylation in genes, pathways, and regulatory transcription factors that link to anti-viral processes. We found that hypermethylated genes were enriched in the regulators of transcription, metabolism, and epigenetic processes. These findings were validated on three publicly available datasets by comparison with seasonal transcriptomic gene modules, seasonal changes in chromatin accessibility, and VDRE/VDR-mediated transcription. Together, our findings add a layer of epigenetic regulation to seasonal changes in immune cell functions, with potential consequences for infectious and inflammatory disease exacerbations.

As we’ve previously described, seasonal immune effects can be mediated by changes in UVB skin exposure and resultant serum vitamin D_3_ levels^9^. In this study, we found that both factors independently change the PBMC methylome, as indicated by shared and distinct Summer and Winter+D_3_ methylome differences in individuals who experienced winter reduced UV exposure and vitamin D supplementation^15^. Although we cannot prove that seasonal changes are a direct result of recorded UVB fluctuations, such epigenetic changes in immune cells are supported by prior observation that UVB exposure of human keratinocytes *in vitro* rewires their methylome^43^, and has been linked to skin cancer progression^44^. Similarly, maternal vitamin D deficiency alters DNA methylation levels in germline and somatic cells in mice^45^. Vitamin D supplements, together with high physical activity (outdoor exercise, a potential correlate of UV exposure) were associated with greater DNA methylation changes in saliva cells from elderly persons (70-95 yrs)^46^. In contrast to our findings, the same study reported only one DMP associated with oral vitamin D supplementation, but this reflected personal multivitamin use, not a standardized vitamin D intervention, and was cross-sectional across a wide age range. The power of our study comes from the longitudinal sampling of the same people of similar age and ancestry, timed to capture seasonal vitamin D peaks and troughs, and after the use of high-dose vitamin D_3_ that achieved optimal serum 25(OH)D levels post-supplementation (median >100nM), thereby reducing biological confounders and variable supplementation use.

We identified TP53 and ESR1 as central upstream transcriptomic regulators of the differentially methylated genes shared between Summer and Winter vitamin D_3_ supplementation. Functionally, increased TP53 activity inhibits HIV-1 infection^47–49^ and, ESR1 signaling blocks HIV-1 reactivation^50^. We observed winter hypermethylation of both *TP53* and *ESR1*, suggesting winter-related downregulation of both genes, aligns with our previous data from the same cohort showing increased HIV-1 replication in Winter compared to Summer PBMC^15^. We also confirmed *ESR1* expression was upregulated in PBMC *in vitro* following 1,25(OH)_2_D_3_ treatment, with a similar trend for *TP53,* although the response was highly variable between donors for both genes, consistent with them also containing DVP.

Transcriptional regulators such as *EP300*, *CREBBP* and *KDM1A* are known modulators of epigenetic changes including DNA methylation^51^ and their identification points to a synergy of complimentary mechanisms resulting in differences in DNA methylation. Differentially methylated and variable genes showed widespread GO enrichment in metabolic processes such as mitochondrial (TCA cycle) and phospholipid homeostasis, epigenetic regulation, cytokine signaling, response to pathogen, oxidative stress, cellular aging, and overall immune response related genes. Supporting the change in DNA methylation with season and by vitamin D_3_, we observed an enrichment of Summer and Winter+D_3_ DMP in one-carbon metabolism related regulators (*MTHFR*, *MTR*, *MTHFD1*) and folate metabolism which are required for DNA methylation changes^52–55^. These genes regulate the S-adenosyl-l-methionine (SAM) methyl donor pool^56,57^, which can be used by DNA-methyl transferases (e.g. DNMT3A) to regulate DNA methylation levels^58^. Seasonal changes in the levels of folate^59^ and serum vitamin D_3_^4^, can also regulate folate availability^59^. Thus, one-carbon metabolism tied to folate and vitamin D_3_ levels might be a unifying mechanism for shared methylation changes^53,59^ between Summer and Winter+D_3_.

Differentially methylated genes both in Summer and Winter+D_3_ were commonly enriched in the PI3K-AKT signaling pathway, with summed z-score indicating overall pathway hypomethylation in Summer and Winter+D_3_. The PI3K-AKT pathway is critical for anti-viral immunity, activating type I Interferons, and is exploited by viruses, such as HIV-1, for spread and has been targeted for HIV cure strategies^60^. Similarly, influenza A virus transiently activates PI3K to gain cell entry but once the host immune system recognizes the virus the PI3K activation is anti-viral^61^ and also promotes cell survival^62^. Relative Winter hypermethylation of this pathway may therefore contribute to reduced ability of circulating immune cells to respond to viral infections. HIV-1 infection is known to impair mitochondrial function, TCA cycle^63^ and phospholipid metabolism^64^, to an extent that anti-phospholipid antibodies inhibit HIV-1 infection of PBMC^65^. Similarly, HIV-1 accelerates immune aging through changes in DNA methylation^66^ and is known to induce oxidative stress^67^. Seasonal and vitamin D-deficiency related perturbations of DNA methylation within these same pathways may therefore contribute to seasonal exacerbations of viral infections, including those in people living with HIV-1^68^. Virus-host interactome analysis revealed shared protein-protein interaction between HIV-1 and SARS-CoV-2 viral proteins and proteins encoded by genes with DMP in Summer and/or Winter vitamin D_3_ supplementation.

Pathway analysis revealed the enrichment of both DMP and DVP in multiple ferroptosis-related genes, which were validated to translate into transcriptional activation by 1,25(OH)D_3_ *in vitro* in uninfected and HIV-1 infected PBMCs. Ferroptosis, an inflammatory programmed cell death pathway, is driven by iron-dependent lipid peroxidation and redox failure, resulting in ROS accumulation^37^. Notably, the top two DMP in Summer and Winter+D_3_, *PLAA* and *COQ5,* which limit ferroptosis, were hypermethylated in winter, suggesting reduced expression and ferroptosis susceptibility. PLAA interacts with and is activated by PDIA3 (previously called 1,25(OH)D_3_-membrane-associated rapid response to steroid), a membrane protein mediating VDR-independent rapid 1,25(OH)D_3_ activity^69^. Our findings highlight multiple layers of vitamin D regulation of *PLAA*, both by modulating its transcription through methylation and by serving as an activating ligand. Validating that 1,25(OH)D_3_ treatment results in increased expression of additional ferroptosis inhibitors, including *GPX4* and *AKR1C1,* points to a broad anti-ferroptotic effect of vitamin D_3_. Such a role is also aligned with vitamin D’s known transcriptional suppression of hepcidin (*HAMP*), resulting in increased ferroportin (*SCL40A1*) expression that exports iron out of a cell^70^. Maintenance of iron homeostasis is important for a variety of infectious diseases, including HIV-1 and tuberculosis^71^. The winter variability in methylation of numerous genes involved in these pathways may therefore contribute to an individual’s seasonal disease exacerbation. A recent study corroborates our observation, which showed that HIV-1 Tat protein induces ferroptosis in microglial cells, down-regulating *GPX4* expression^72^ which we now show in PBMC can be reversed by 1,25(OH)D_3_ treatment. Moreover, ferroptosis has also recently been identified to be seasonally epigenetically regulated at the level of accessible chromatin in PBMC from individuals following BCG vaccination^73^.

The strength of our study is the longitudinal sampling of individuals within tight windows, timed to match local environmental events. The significance of our longitudinal findings also emphasizes the importance of correcting for differences in sample collection time points between individuals in cohort studies, particularly when recruiting in regions of high latitude that have the largest seasonal UV fluctuations. Our findings provide insight into seasonal variations of the DNA methylome that are consistent with seasonal blood transcriptome identified in individuals living at large distances from the equator, where seasonal UV fluctuations are most extreme^10^. Thus, our data support broad epigenetic underpinning of documented seasonal transcriptomic oscillations.

Our study has some limitations. Although, we validated our findings across a variety of publicly available datasets, direct *ex vivo* PBMC transcriptomic data was not available for validation of our methylome findings using matched sample. While pooling the DNA for assessing DNA methylation helped us identify median differences, a previously validated approach^74^, this would have masked some inter-individual variation. Therefore, validation of these results in additional longitudinal cohorts and in populations with different environmental exposures and genetic backgrounds will further strengthen the findings and determine the generalizability as well as population-specific nature of the changes observed.

In conclusion, we identified seasonal methylome changes in PBMC of healthy young adults that differ between summer and winter and are linked to cellular processes affecting metabolic state, antioxidant response, immune function and anti-viral immunity. By assessing the modulation of these pathways after six-weeks of high-dose oral vitamin D_3_, we confirmed the seasonal epigenetic components directly modified by vitamin D_3,_ as well as season-independent vitamin D_3_ effects. Our findings support the idea that maintaining long-term sufficient vitamin D_3_ levels through supplementation could support anti-viral immunity in those at risk of seasonal deficiency. Our findings further demonstrate that analyzing the steady-state methylome can identify biological processes associated with the outcome of infection of those cells and shed light on underexplored mechanisms of disease pathogenesis.

## MATERIALS AND METHODS

### Cohort and study design

The cohort was enrolled in Cape Town, South Africa and study inclusion criteria and phlebotomy has been described previously^15^. The study was ethically conducted according to the Helsinki 1964 declaration and approval received from the Human Research Ethical Committee of the Faculty of Health Sciences, University of Cape Town (ref. 003/2013 and 317/2016). For participants undergoing longitudinal blood collection, informed written consent was obtained for the study as previously described^15^. Stored PBMC from 30 HIV-1-uninfected individuals (13 females, 17 males) 18-24 years of self-reported Xhosa ancestry were selected. These individuals had PBMC available at three time points: summer, winter and winter after 50,000 IU of cholecalciferol (vitamin D_3_)/week for 6-weeks. For *in vitro* validation experiments PBMC were isolated from fresh buffy coats from 5 healthy adult donors collected in winter from the Western Cape Province Blood Transfusion Service, Cape Town, South Africa.

### Whole blood immunophenotyping for absolute cell counts

Whole blood (WB) absolute immune cell counts were determined by flow cytometry using differential leukocyte counting and immunophenotyping in cryopreserved *ex vivo* whole blood (DLC-ICE)^75^, staining lineage-defining cell surface markers for neutrophils, monocytes, T cells, B cells, natural Killer (NK) cells, myeloid dendritic cells (mDC), and plasmacytoid dendritic cells (pDC) (**Suppl. Table S2**). Briefly, WB was fixed in 1X FACS Lyse solution (BD) for 15 min, centrifuged at 2100 X g for 7 min at room temperature and the cell pellet resuspended in cryosolution (50%RPMI (Gibco), 40% Fetal bovine serum (FBS; Thermo Fisher Scientific) and 10% dimethyl sulfoxide (DMSO; Sigma Aldrich)) and aliquots stored at −80°C overnight in a CoolCell (Corning) before transfer to liquid nitrogen for long-term storage. Frozen cells were thawed, washed, permeabilized (1X Perm/Wash (BD)) and stained as previously described^75^ using a 12-colour antibody cocktail (**Suppl. Table S2**) diluted in 50 μL of Brilliant Stain Buffer (BD) for 45 min at 4°C and resuspended in 100 μL PBS after washing in 1X Perm/Wash. Single-stained controls were prepared for each antibody and run with each acquisition. Flow-Count Fluorospheres (Beckman Coulter, 50 μL) were added to each participant sample before acquisition and samples acquired on a BD LSRII flow cytometer with 4 lasers (488, 405, 635, and 532 nm). FCS files were imported into FlowJo software (BD, version 9) to compensate and analyze data. Example gating strategy for identifying immune cell lineages is shown in **Suppl. Fig. 1a**. Event counts of each cell subset and Flow-Count beads were exported and absolute cell numbers/μL of blood were calculated as described^75^. Cell counts were analyzed between visits by Friedman with Dunn’s multiple comparison test.

### DNA isolation, pooling, and methylation evaluation

DNA was isolated from stored PBMC using the DNeasy Blood and tissue Kit from QIAGEN, according to the manufacturer’s instruction. Since it has been shown for methylation analysis that DNA pooling identifies similar number of differential probes as single samples^74^, we decided to use this approach. Pooling at least six samples has been suggested to be a good approach, and we followed the same^76^. From the 30 individuals, DNA from the same six individuals were pooled at each of the three times, creating five pools per time point and a total of 15 pools (**Fig. 1a**). Double stranded DNA concentration was determined using the Qubit fluorometric test (Invitrogen). At least 500 ng of DNA was bisulphite converted and used as input on the Illumina Infinium EPIC DNA methylation array (v1.0) and post hybridization the chip was scanned using the iScan system (Illumina). Processing of Illumina chips was conducted by the Clinical Genomic Center, Toronto, Ontario, Canada.

### Differential methylation and variability analyses

Epic array raw data IDAT files were processed using the *minfi* package in R. The data was normalized using *PreprocessQuantile* normalization. The data was visualized using PCA using the *prcomp* package in R. Data were plotted for filtered probes using a M-value standard deviation of at least 0.1 then a more restrictive filtering using standard deviation > log_2_ 1.25, after regressing out the donor pool effect. Differential methylation was analyzed using *limma* to identify a differential probe on the normalized M-values with > 0.1 standard deviation. We used the moderated variance-based F-test as implemented in *limma* R package to test for differential methylation. Akin to conventional ANOVA, it tests whether there is an overall difference in methylation at a per-probe level, irrespective of the group being tested. The resulting two-sided p-values were adjusted for multiple testing using storey’s method of *pi0.est* estimation as implemented in the *siggenes* package in R. For the F-test, Winter was used as the comparator against Summer (S-W), and Winter+D_3_ (D-W), the difference between these two comparisons was also analyzed (D-S). Top DMP were assigned applying a false discovery rate (FDR, q < 0.25) and post-hoc pairwise significance of two-sided p < 0.01 for at least on timepoint comparison. In differential methylation analysis, due to methylation variability between samples within the same comparison group, the discovery of significant differential methylation using pair-wise comparisons is hindered; in such cases differential variability analysis has been shown to discover differences between groups^17^. Differential variability of CpG methylation was assessed on M-values using the *DiffVar method* implemented in the *missMethyl* package in R, with top DVP assigned at the same FDR and a stricter timepoint comparison using two-sided p < 0.001. Significance of probe Beta-value distribution between time points for all raw, normalized and Top DMP was analyzed by two-sample KS test with two-sided p-values for each apir of seasons (S vs W, S vs D, W vs D). Ill

We used the methylation directionality and paired genomic location to define ‘Gene-activating’ DVP/DMP based on CpG sites that were either hypomethylated in Summer or Winter+D_3_ and annotated as TSS1500, TSS200, 5ʹ-UTR or 1stExon on the array annotation manifest; or hypermethylated and annotated as Body or 3ʹ-UTR. Accordingly, ‘gene-repressing’ DMP/DVP were defined as CpGs that showed hypermethylation at gene promoters or hypomethylation at gene bodies^17^. Genomic region enrichment was tested by hypergeometric over-representation test, with the background (“universe”) defined as all probes that passed QC and SD filtering. P-values from the hypergeometric tests were corrected for multiple testing (FDR), and categories with FDR < 0.05 considered significantly enriched. Enrichment scores shown in the figures correspond to the log₂(observed/expected) ratio for each category (positive = over-represented, negative = under-represented). Separate analyses were run for hypermethylated (FC >0 as compared to winter), hypomethylated DMPs (FC < 0 as compared to winter), and similarly for DVPs.

### Pathway enrichment, regulator and network analysis

Upstream regulator analyses were performed in Ingenuity Pathway Analysis (IPA, QIAGEN) using methylation-derived gene lists. For DMPs, we used the TopF set (q < 0.25 and p < 0.01), for DVPs, we used genes corresponding to CpGs with DiffVar q < 0.25. These gene lists were uploaded as “focus molecules” and analyzed with IPA’s Upstream Regulator Analysis. The reference set was the Ingenuity Knowledge Base (Genes Only); both direct and indirect relationships were allowed; species was restricted to human; confidence was set to Experimentally observed; tissue was set to All tissues; and all molecule types and databases were included. IPA calculated an overlap p-value for each regulator using Fisher’s exact test. Regulators with p < 0.05 were retained, and we further filtered to regulators with ≥ 5 downstream target genes in the input list for interpretation and plotting. This was extracted and the list used in the STRING protein-protein interaction database. Only experimentally verified interactions were considered. This interactome was visualized using Gephi, which computed the hub and centrality measures. The graph was then exported to Cytoscape for final visualization and presentation. For pathway-level comparisons, the core analysis as run above using the same settings for DMP and DVP were entered into IPA for comparison analysis. This analysis compares canonical pathways across multiple lists based on shared pathway genes. The output is an overlap p-value list across comparisons, indicating whether the pathways were unique or shared. Once obtained, the pathways were plotted and common themes identified using shared reactome terms.

Gene ontology analysis was carried out using the *goseq* package using the *go2sy* using GO:CC, GO: BP, GO:MF and KEGG processes and an overrepresentation p-value derived using the *pi0.est* estimation. *Goseq* explicitly corrects for gene-length / probe-count bias by estimating a probability weighting function from the number of EPIC CpG probes mapped to each gene and its DE status and then computing bias-adjusted over-representation p-values. The input lists were from the F-test derived differential lists with a cutoff of p < 0.01 for DMP and the *DiffVar* lists with p < 0.001 for DVP. Any enriched processes with goseq p < 0.05 were considered significant. A similar analysis for DMP was carried out using the C2 module of the Molecular Signatures database (MsigDB). The GO terms were merged into consolidated terms based on shared Reactome definitions, which were curated manually for each enriched ontology to assign the composite/consolidated pathway terms. Methylation beta values of the genes enriched in significantly overrepresented consolidated pathways were used to calculate a per gene z-score per sample in R and visualized using box plots generated in Prism (GraphPad v10.4.1).

Initial GO analysis yielded more than 3000 terms which passed a cut-off of p <0.05, to obtain a more tractable result, the obtained List of GO terms was separated based on the ontology category (GO:CC, GO: BP, GO:MF) and sorted by the most significant p-values and the highest gene set size (the number of enriched genes). The top 50% of the total GO terms obtained from each list were compiled in a combined list of GO terms. This list was visualized using *Enrichment map* in Cytoscape (Supplementary Fig. S3). The network connection is based on the shared genes among the GO terms, represented as the edge (line connecting two nodes), and the node is each GO term. Nodes having multiple edges would mean that GO terms have the highest gene set size and thus have multiple shared genes with other GO terms. *AutoAnnotate* function within *Enrichment Map* was used to automatically interpret and merge the terms to get overall impressions, represented as boxes and terms above them. *AutoAnnotate*, automatically generates cluster labels based on word frequencies of selected attributes^77^. In case of ambiguous terms, it was replaced by the GO term with the highest p-value in the Enrichment Map collection. The *Enrichment map* enriched q-value was plotted in the composite Figure.

Metabolic and Epigenetic regulators were identified from the genes that mapped to the Metabolic processes, and Epigenetic regulation. DVPs were enriched in more of these processes, so this analysis focused on the pathways and genes from DVPs. Within the pathway, we classified the enzymes based on their specific metabolic process and rate limiting enzyme classification based on the RLEdb^23^. Methylation beta values were used for plotting the heatmaps and z-score of normalized scores per-gene is presented in the heatmaps. Since the values come from DVPs, we refer to them here as methylation variability. The values extracted as a table were plotted using *complexheatmap* package in R.

### HIV-1 and SARS-CoV-2 interactome

The HIV-1 and human protein-protein interactome was downloaded from the NIH database^28^ and the SARS-CoV-2-human protein-protein interactome from Gordon et al.^32^. The top summer and Winter+D_3_ DMP list were checked for overlaps using a Venn for each viral interaction separately. The overlapping genes were exported to a list and used as an input in Gephi 0.92^78^ to visualize the network for each virus. The Force Atlas 2 visualization^79^ was used to obtain the presented network. The network was exported as a network file, which was imported for the final visualization in Cytoscape21.

### Orthogonal validation datasets

Three external validation datasets were used. The first seasonal dataset gene list was obtained from supplementary materials of Dopico et al.^10^. The data from UCAR seasonal chromatin accessibility^12^ was kindly provided by Dr. Dyugu Ucar after email correspondence and consisted of the genomic co-ordinates of the differential season peaks (8744 peaks). This was then mapped to 3638 unique genes using PAVIS^80^ and this list was used for overlap analysis as shown in **Fig. 5c**. The VDR Chip Seq data and the Vitamin D regulated gene data were available as supplementary tables in Tavera-Mendoza et al.^14^.

### PBMC HIV-1 infection

HIV-1 stocks were prepared by using HIV-1 strain Ba-L which was grown in PBMC isolated from healthy donor buffy coats in RPMI containing 50 mM glutamine, 20% (vol/vol) fetal calf serum (FCS), 100 IU/mL penicillin, 100 g/mL streptomycin, and 20 IU/mL IL-2 (Sigma), as previously described^15^. Culture supernatants were first passed through a 0.22-uM PVDF filter (Millipore) to remove cellular debris and then purified before use by ultracentrifugation through a 20% (wt/vol) sucrose buffer and resuspended in RPMI-1640 medium with 5% (vol/vol) human AB (HAB) serum (Invitrogen). HIV-1 virus preparations TCID50 were calculated by titration on TZM-bl cells, and following cell lysis in BrightGlo (Promega) luminescence measured using a GloMax-96 (Promega).

For gene expression experiments following HIV-1 infection, PBMC were suspended at 1 × 10^7^ cells/mL in RPMI-1640 medium with 10% (vol/vol) HAB serum. Then, 150 μL aliquots were mixed with 30 μL aliquots of purified HIV-1 Ba-L [MOI 0.004] or RPMI with 5% HAB serum (vol/vol) (infection control), plated in triplicate in a U-bottom 96-well plates (60 μL per well), and incubated for 17 hours at 37 °C at 5% CO2. After incubation, cells were washed three times in warm RPMI before being resuspended for 9 days in 200 μL RPMI with 10% HAB, with and without 100 nM 1,25(OH)D_3_. After 3, 6, and 9 days, half vol (100 μL) of supernatant was removed before adding fresh medium according to the conditions and 1,25(OH)D_3_ supplementation. The PBMC at end of Day 9 were lifted from the 96-well plate using 20mM EDTA, combining the cells from the triplicate and stored in TRIzol (Invitrogen) at −80°C for further use.

### Quantification of the gene expression by qPCR

RNA was extracted from TRIzol samples using RNeasy kits (Qiagen) and precipitated with isopropanol (Sigma-Aldrich), sodium acetate (Sigma-Aldrich), and linear polyacrylamide (Life Technologies) at 20 °C overnight. Before final resuspension in RNAase free H_2_O, the precipitated RNA was washed twice with 80% ethanol (Sigma-Aldrich). 100-200 ng RNA from the PBMC was reverse transcribed using the High-Capacity cDNA synthesis kit (Applied Biosystems), and real-time RT-PCR was performed on the Quantstudio 7 (Applied Biosystems) with FAST SYBR green, 10% cDNA, 60 °C annealing temperature, and 40 cycles. To confirm amplicon specificity, melt curve analysis was performed. Relative quantification was performed using synthesized primers (Sigma-Aldrich) which were designed in house listed here: *COQ5* FP: 5’ CCGTCTCAGGAAGAGTTCAAGG 3’, *COQ5* RP: 5’ TTCCAGGCTTTCAACAGGATATG 3’; *PLAA* FP: 5’ TGTGGTTGGTGCGAGTGATG 3’, *PLAA* RP: 5’ AATCAATGGTTGCGTGAGACAG 3’; *TP53* FP: 5’ CCTGAGGTTGGCTCTGACTGTAC 3’, *TP53* RP: 5’ CCAGTGTGATGATGGTGAGGATG 3’; *ESR1* FP: 5’ CCCACATCAGGCACATGAGTAAC 3’, *ESR1* RP: 5’ CGTCCAGCATCTCCAGCAG 3’; *HMOX1* FP: 5’ CTGCTGACCCATGACACCAAG 3’, *HMOX1* RP: 5’ AGTGTAAGGACCCATCGGAGAAG 3’; *ATG5* FP: 5’ CGAGATGTGTGGTTTGGACG 3’, *ATG5* RP: 5’ ACTCTTGGCAAAAGCAAATAGTATG 3’; GPX4 FP: 5’TCGGGAAGCAGGAGCCAG 3’, *GPX4* RP: 5’ TCTTCATCCACTTCCACAGCG 3’; *AKR1C1* FP: 5’ CAATTGAAGCTGGCTTCCG 3’, *AKR1C1* RP: 5’ TCTGGTCGATGGGAATTGCA 3’. *RPL13A* was used as the housekeeping gene and CYP24A1 as a 1,25(OH)D_3_ positive control using primers previously described^81^. Primers were tested for amplification efficiency by using a standard curve on a serially diluted cDNA and primers with a slope close to –3.3 were considered good primers to use. The cDNA from our experimental conditions were also tested to ensure they were in detection range of the standard curve. Gene Ct values were normalized to the housekeeping gene RPL13A and fold change for each experimental conditions for each donor compared to their corresponding uninfected control values using 2-ɅλɅλCT method. Fold change values were analysed by Freidman test with Benjamini, Krieger and Yekutieli False discovery rate.

## Acknowledgements

We would like to thank all participants for volunteering to participate in the study, Dr Guillaume Borque for helpful discussion during initial stages of methylome analysis, and Dr. Dyugu Ucar for providing the seasonal chromatin accessibility data upon request. The work was supported by funding from AXA Research Fund (Grant no: 25776), National research Foundation, South Africa (UID: 88297), Academy of Science of South Africa, John Simon Guggenheim Fellowship, the Stellenbosch Institute for Advanced Study, and Wellcome (203135Z/16/Z and 226817). RJW is supported by the Francis Crick institute which receives funding from Wellcome (CC2112), Cancer Research UK (CC2112) and the Medical Research Council (CC2112). He also receives support in part from the NIHR Biomedical research Centre of Imperial College Healthcare NHS Trust. The funders had no role in study design, data collection and analysis, decision to publish, or preparation of the manuscript. For the purposes of open access, the authors have applied a CC-BY public copyright to any author-accepted manuscript arising from this submission.

## Author Contributions

A., A.K.C., and R.J.W. designed the epigenetics study. A.K.C, R.J.W., and N.G.J. designed and collected cohort samples. A. and L.V. performed bioinformatic analyses with input from A.K.C. A.K.C and E.N. designed and performed flow cytometry analysis, A. and A.K.C. designed in vitro experiment, performed and analyzed by A. Manuscript was drafted by A. and A.K.C and edited by all authors.

## Data Availability

The methylation data generated for the paper has been deposited to the GEO, accession number: GSE303851. The data will be made publicly available upon acceptance of the manuscript.

