## Supplementary Figures for "Seasonal and Vitamin D3 mediated methylome reprogramming of peripheral immune cells and anti-viral responses"

[1] Wellcome Discovery Research Platforms in Infection, Institute of Infectious Disease and Molecular Medicine, Departments of Pathology and Medicine, University of Cape Town, Observatory, South Africa. [2] Department of Microbiology, Biological Sciences Division, The University of Chicago, Chicago, Illinois, USA. [3] Canadian Centre for Computational Genomics (C3G) McGill University, Montreal, QC, Canada. [4] Department of Human Genetics, McGill University, Montreal, QC, Canada. [5] Victor Phillip Dadelahi Institute of Genomic Medicine, McGill University, Montreal, QC, Canada. [6]South African Tuberculosis Vaccine Initiative, Division of Immunology, Department of Pathology and Institute of Infectious Disease and Molecular Medicine, University of Cape Town, Cape Town, South Africa. [7]Department of Anthropology, The Pennsylvania State University, State College, United States. [8] Department of Infectious Diseases, Imperial College London, London, United Kingdom. [9]The Francis Crick Institute, London, United Kingdom. [10]Walter and Eliza Hall Institute of Medical Research, Parkville, VIC, Australia. [11]Division of Medical Biology, Faculty of Medicine, Dentistry and Health Sciences, University of Melbourne, Melbourne, VIC, Australia.

*Corresponding authors

Anna Coussens,

Abhimanyu,

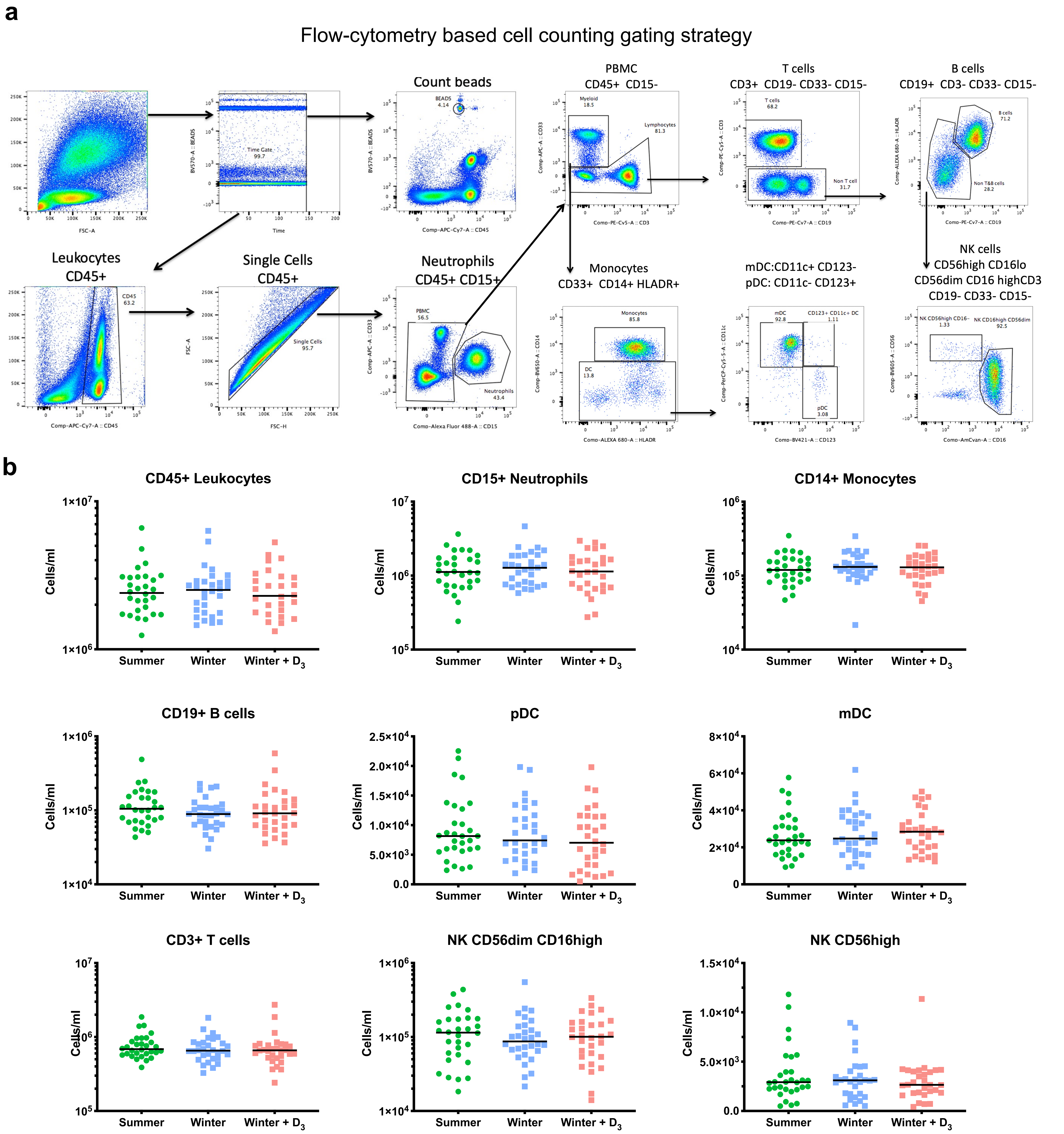


**Figure S1. Longitudinal immune cell population counts. a)** Flow cytometry gating and markers used for ascertaining absolute cell counts for each population shown in **b)** presented as median cell count/ml blood, n=30 per time point. No significant differences were identified between time points. Friedmann Test with Dunn's post-hoc correction. MN: monocytes, mDC: myeloid Dendritic cells, pDC: plasmacytoid dendritic cells, NK: Natural killer cells.

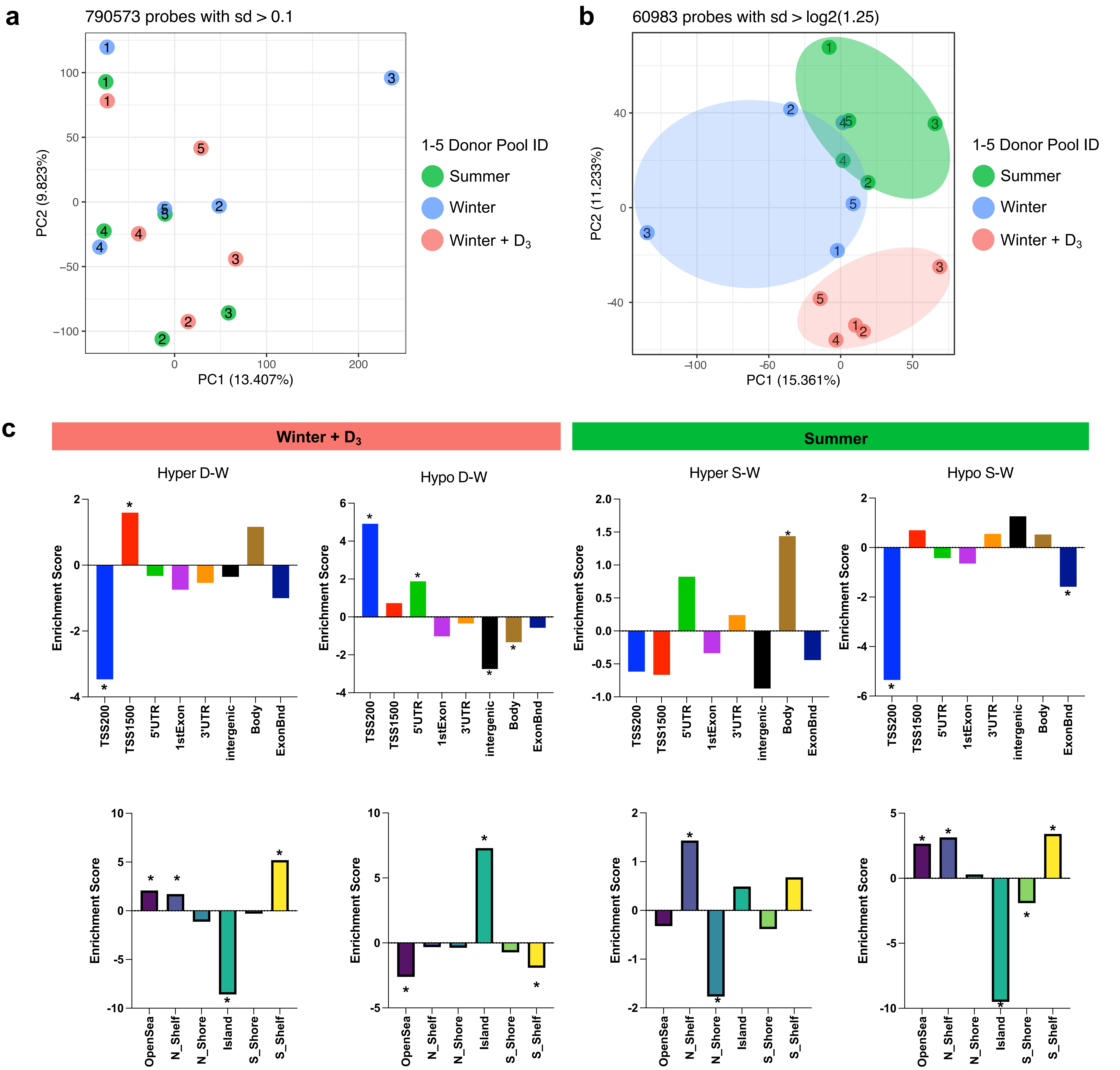


**Figure S2. Genome-wide methylation sample variability and genomic distribution.** **a)** M-value principal component analysis (PCA) plot of all 790,573 CpG probes showing standard deviation of >0.1; **b)** PCA plot of probes after regressing out the pool effect and filtering on probes with SD > log21.25, samples cluster by timepoint. The circle colors indicate the three timepoints and numbers in the circle indicate the DNA pool that the sample belongs to; **c**) The enrichment scores for the genomic and CpG location of the significant differentially methylated probes, the whole list is supplied as Table S3. The significant enrichment is shown with an Asterix (*).

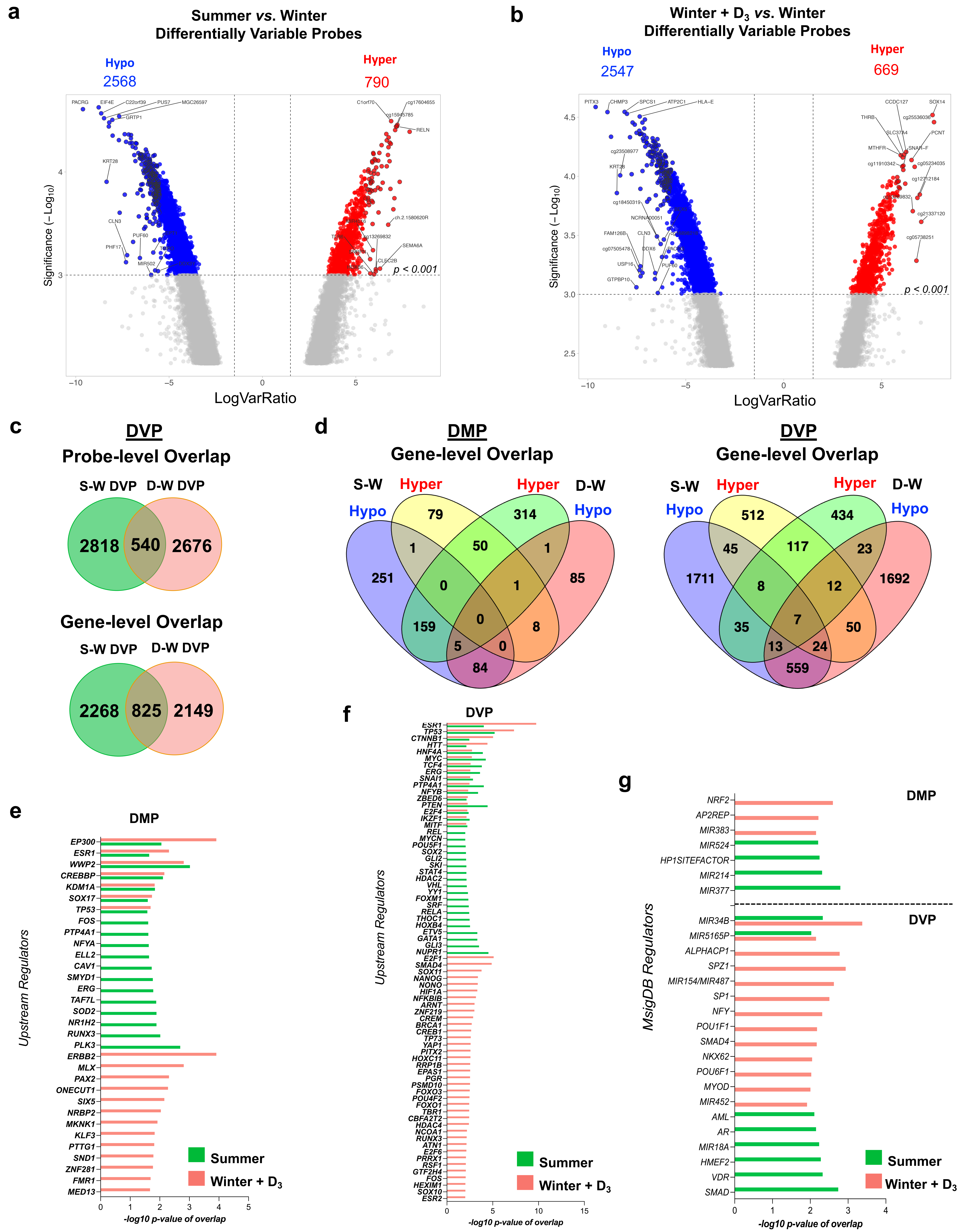

**Figure S3. Shared and unique Differential features and predicted Transcription regulators using Ingenuity pathway analysis and MSigDB**. Volcano plot of differentially variable probes (DVP) in (**a**) Summer, and (**b**) Winter+$D_3$; (**c**) Venn diagram of DVP at the probe and gene level; (**d**) Venn diagram of differentially methylated probes (DMP) and DVP at the gene-level indicating the number of shared hyper- and hypo-methylated genes for Summer (S-W), and Winter+$D_3$ (D-W), compared to Winter; **e)** Overlapping and unique top predicted transcription factors (TF) for differentially methylated probes**; f)** Overlapping and unique top predicted TF for differentially variable probes; **g)** Overlapping and unique TF for DMP and DVP as predicted by MsigDB.

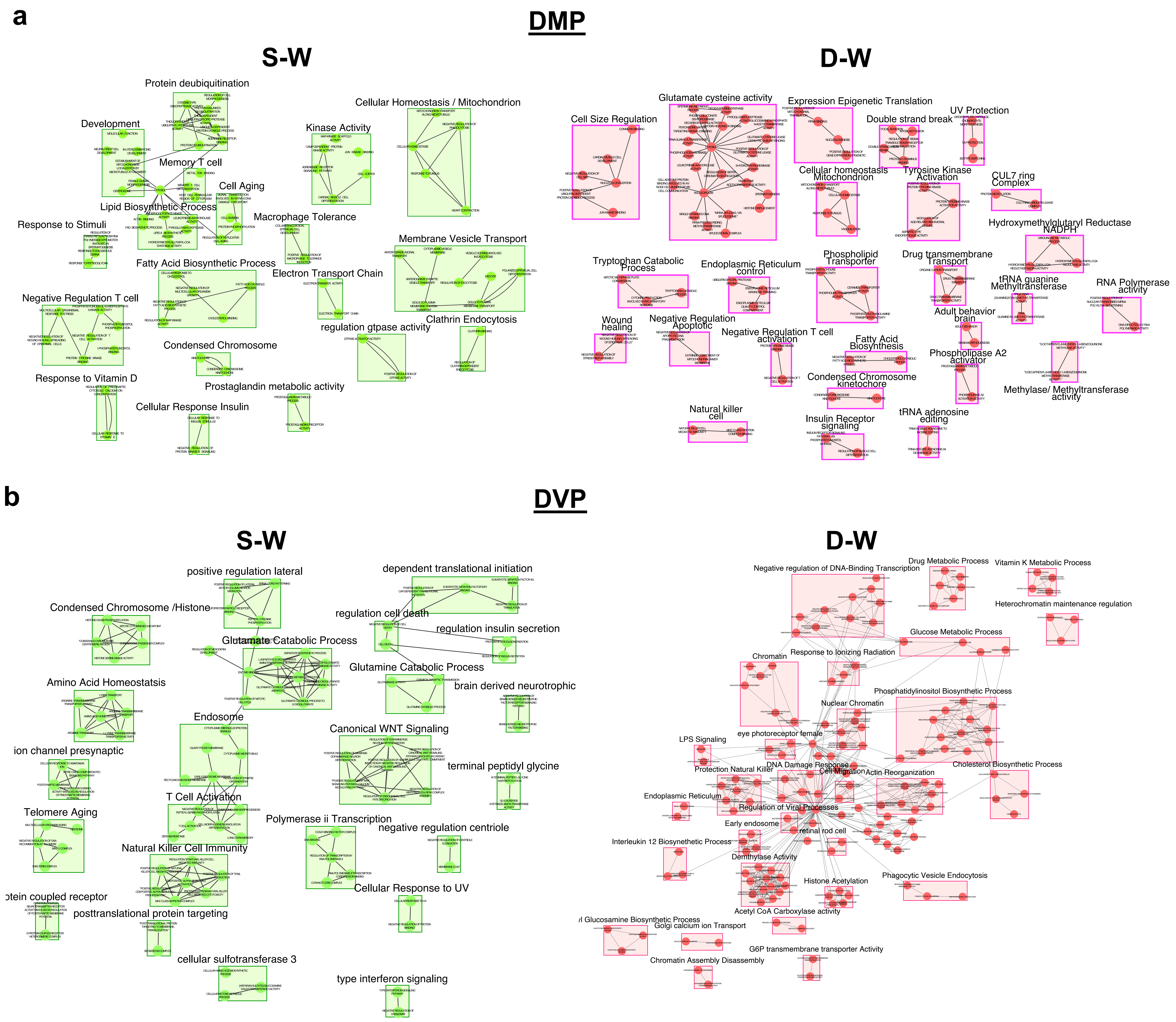


**Figure S4. Enrichment map of shared GO terms summarized in Fig. 3a.** For **(a)** differentially methylated probes (DMP) and **(b)** differentially variable probes (DVP) for Summer vs Winter (S-W) and Winter+$D_3$ vs Winter (D-W). Each node is a GO term as derived from a hypergeometric test and each edge (connection) shows that the GO terms have similar and shared genes. Each such closely related terms are then subject to enrichment using enrichment map and similar terms are put in a box and a consensus name generated by the software with a significant q value of overlap.
